# Random glucose GWAS in 493,036 individuals provides insights into diabetes pathophysiology, complications and treatment stratification

**DOI:** 10.1101/2021.04.17.21255471

**Authors:** Vasiliki Lagou, Longda Jiang, Anna Ulrich, Liudmila Zudina, Karla Sofia Gutiérrez González, Zhanna Balkhiyarova, Alessia Faggian, Shiqian Chen, Petar Todorov, Sodbo Sharapov, Alessia David, Letizia Marullo, Reedik Mägi, Roxana-Maria Rujan, Emma Ahlqvist, Gudmar Thorleifsson, He Gao, Evangelos Evangelou, Beben Benyamin, Robert Scott, Aaron Isaacs, Jing Hua Zhao, Sara M Willems, Toby Johnson, Christian Gieger, Harald Grallert, Christa Meisinger, Martina Müller-Nurasyid, Rona J Strawbridge, Anuj Goel, Denis Rybin, Eva Albrecht, Anne U Jackson, Heather M Stringham, Ivan R Corrêa, Farber-Eber Eric, Valgerdur Steinthorsdottir, André G Uitterlinden, Patricia B Munroe, Morris J Brown, Schmidberger Julian, Oddgeir Holmen, Barbara Thorand, Kristian Hveem, Tom Wilsgaard, Karen L Mohlke, Wolfgang Kratzer, Haenle Mark, Wolfgang Koenig, Bernhard O Boehm, Tricia M Tan, Alejandra Tomas, Victoria Salem, Inês Barroso, Jaakko Tuomilehto, Michael Boehnke, Jose C Florez, Anders Hamsten, Hugh Watkins, Inger Njølstad, H-Erich Wichmann, Mark J Caulfield, Kay-Tee Khaw, Cornelia van Duijn, Albert Hofman, Nicholas J Wareham, Claudia Langenberg, John B Whitfield, Nicholas G Martin, Grant Montgomery, Chiara Scapoli, Ioanna Tzoulaki, Paul Elliott, Unnur Thorsteinsdottir, Kari Stefansson, Evan L Brittain, Mark I McCarthy, Philippe Froguel, Patrick M Sexton, Denise Wootten, Leif Groop, Josée Dupuis, James B Meigs, Giuseppe Deganutti, Ayse Demirkan, Tune H Pers, Christopher A Reynolds, Yurii S Aulchenko, Marika A Kaakinen, Ben Jones, Inga Prokopenko, on behalf of the Meta-Analysis of Glucose and Insulin-related Traits Consortium (MAGIC)

**Author notes:** These authors contributed equally to this research (Vasiliki Lagou, Longda Jiang, Anna Ulrich). Genentech, 1 DNA Way, South San Francisco, CA 94080. These authors jointly directed this research. Correspondence should be addressed to: Prof Inga Prokopenko, Section of Statistical Multi-Omics, Department of Clinical and Experimental Medicine, University of Surrey Leggett Building, Daphne Jackson Road, Manor Campus, Guildford, Surrey, UK, GU2 7WG; Dr Ben Jones, Section of Endocrinology and Investigative Medicine Imperial College London, Du Cane Road London, UK, W12 0NN; Dr Marika Kaakinen, Section of Statistical Multi-Omics, Department of Clinical and Experimental Medicine, University of Surrey Leggett Building, Daphne Jackson Road, Manor Campus, Guildford, Surrey, UK, GU2 7WG.

## Abstract

Homeostatic control of blood glucose requires different physiological responses in the fasting and post-prandial states. We reasoned that glucose measurements under non-standardised conditions (random glucose; RG) may capture diverse glucoregulatory processes more effectively than previous genome-wide association studies (GWAS) of fasting glycaemia or after standardised glucose loads. Through GWAS meta-analysis of RG in 493,036 individuals without diabetes of diverse ethnicities we identified 128 associated loci represented by 162 distinct signals, including 14 with sex-dimorphic effects, 9 discovered through trans-ethnic analysis, and 70 novel signals for glycaemic traits. Novel RG loci were particularly enriched in expression in the ileum and colon, indicating a prominent role for the gastrointestinal tract in the control of blood glucose. Functional studies and molecular dynamics simulations of coding variants of *GLP1R*, a well-established type 2 diabetes treatment target, provided a genetic framework for optimal selection of GLP-1R agonist therapy. We also provided new evidence from Mendelian randomisation that lung function is modulated by blood glucose and that pulmonary dysfunction is a diabetes complication. Thus, our approach based on RG GWAS provided wide-ranging insights into the biology of glucose regulation, diabetes complications and the potential for treatment stratification.

## Main text

Genetic factors are important determinants of glucose homeostasis and type 2 diabetes (T2D) susceptibility. Heritability of both fasting glucose (FG) and T2D is high, at 35-40%^1^ and 30-60%^2^, respectively. To date, more than 400 genetic loci have been described for T2D^3, 4^. Genome-wide association studies (GWAS) for glycaemic traits in individuals without diabetes have identified genetic predictors of blood glucose, insulin and other metabolic responses during fasting or after oral or intravenous glucose challenge tests^5–8^. However, physiological glucose regulation involves responses to diverse nutritional and other stimuli that were, by design, omitted from such studies. Blood glucose is frequently measured at different times throughout the day in clinical practice and research studies (random glucose; RG). Whilst RG is inherently more variable than standardised measures, we reasoned that, across a very large number of individuals, it may more comprehensively represent complex glucoregulatory processes occurring in different organ systems. Therefore, to identify and functionally validate genetic effects influencing RG, explore its relationships with other traits and diseases, and utilise these data to inform approaches to T2D treatment stratification, we performed the first large-scale trans-ethnic GWAS meta-analysis for RG in individuals without diabetes.

### RG GWAS significantly expands the catalogue of glycaemia-related genetic associations

We undertook RG GWAS in 493,036 individuals without diabetes of European (n=479,482) and other ethnic (n=16,554) descent with adjustment for age, sex and time since last meal (where available), along with exclusion of extreme hyperglycaemia (RG>20 mmol/L) and individuals with diabetes (**Supplementary Table 1)**. The covariate selection was done upon extensive phenotype modelling **(Methods, Supplementary Table 2, Supplementary Figure 1a).** We identified 162 distinct signals (*P*<10^−5^) within 128 genetic loci reaching genome-wide significance (*P*<5×10^−8^) (**Figure 1a**, **Supplementary Table 3**). Seventy RG signals had not previously been reported for glycaemic traits (**Table 1, Supplementary Table 3**). In Europeans, while the UK Biobank (UKBB) study provided 83.8% of the total study size, 128 detected signals out of the 143 were directionally consistent in UKBB and other contributing studies grouped together (**Supplementary Table 3**). Adjustment for last meal timing (**Supplementary Figure 1b**) reduced effect sizes for several loci, including *ITPR3*, *RREB1*, *RGS17*, *RFX6*/*VGLL2* and *SYNGAP1*, suggesting that these may be more related to the post-prandial state. *RREB1*, *RFX6* are transcription factors implicated in the development and function of pancreatic beta cells^9, 10^, and *ITPR3* is a calcium channel involved in islet calcium dynamics in response to glucose and G protein-coupled receptor (GPCR) activation^11^. Neither adjustment for body-mass index (BMI), nor a more stringent hyperglycaemia cut-off (RG>11.1 mmol/L or HbA1c≥6.5%) (**Supplementary Figure 1c-e**) materially changed the magnitude and significance of the RG effect estimates, although when all covariate models were individually applied, nine additional signals at genome-wide significance were identified in UKBB (**Table 1, Supplementary Table 4**).

**Figure 1.**
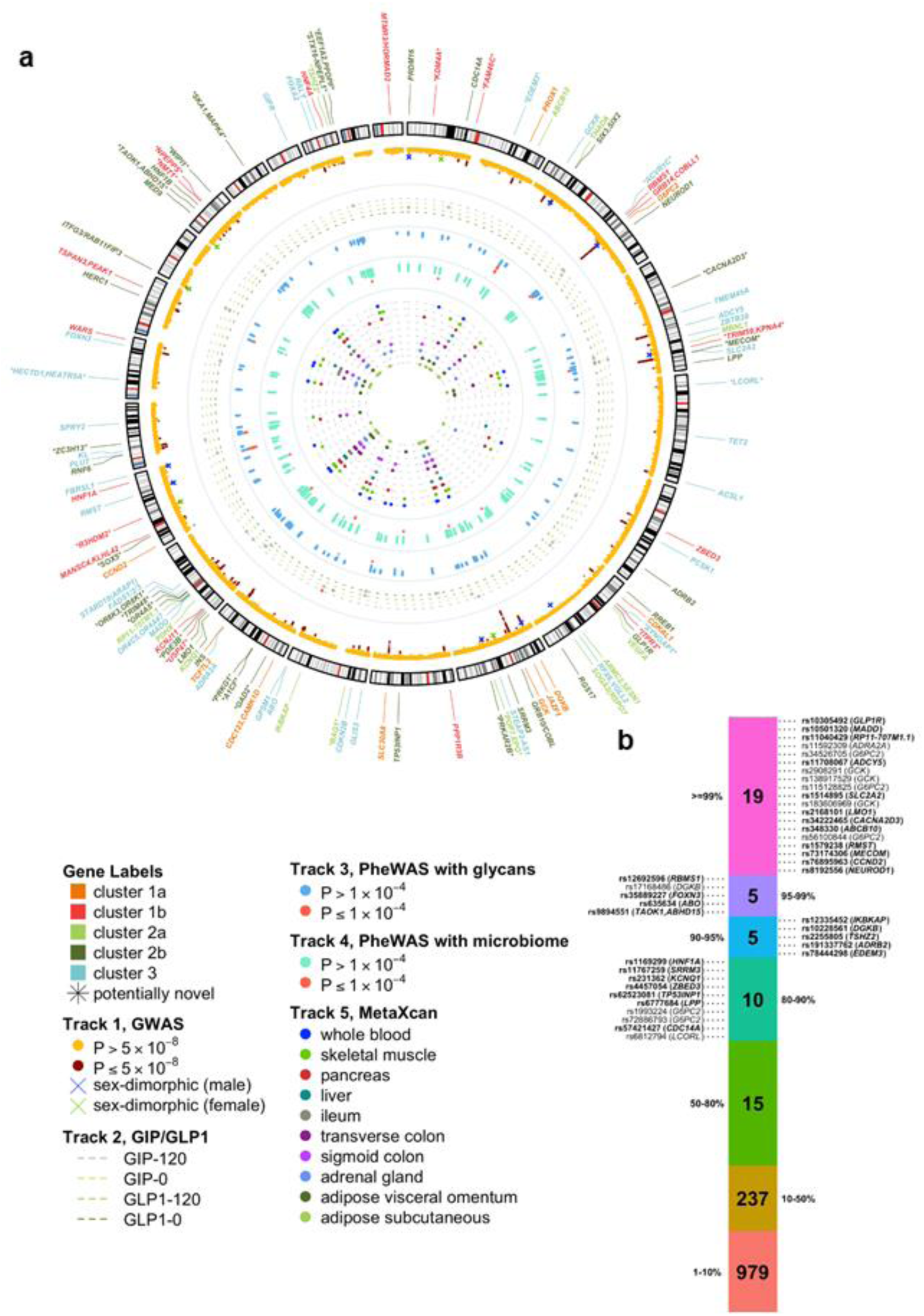
Summary of all RG loci identified in this study. **(a) Circular Manhattan plot summarising findings from the present study. Outermost layer:** Gene names of the 162 distinct RG signals are labelled with different colours indicating three clusters defined in cluster analysis: 1a/b=metabolic syndrome, 2a/b=insulin release *versus* insulin action (with additional effects on inflammatory bowel disease for cluster 2a), 3=defects of insulin secretion (**Methods**). Asterisks annotate novel for glycaemic traits RG signals. **Track 1:** RG Manhattan plot reporting −log10(*P-*value) for RG-GWAS meta-analysis, signals reaching genome-wide significance (*P-*value<5×10^−8^) are coloured in red. Crosses annotate genome-wide significant loci that show evidence of sex heterogeneity (**Methods**): blue crosses indicate signals with larger effects in men, green crosses – signals with larger effects in women. **Track 2:** Effects of RG genome-wide significant on four GIP/GLP-1-related traits GWAS. The colours of the dotted lines indicate four GIP/GLP-1-related traits, grey dot - signals reaching *P-*value<0.01 for a GIP/GLP-1-related trait, red dot – lead SNP has significant effect on GIP/GLP-1-related trait (Bonferroni-corrected *P-*value<1×10^−4^). **Track 3:** Effects [-log10(*P-*value)] of lead RG variants in 113 glycan PheWAS. Blue dots - RG lead SNPs, red dots - lead SNPs reaching *P*-value<10^−4^. **Track 4:** Effects [-log10(*P-*value)] of lead RG variants in 204 gut-microbiome PheWAS. Light green dots - RG lead SNPs, red dots - variants with significant effects at *P*-value<10^−4^. **Track 5:** MetaXcan results for 10 selected tissues for RG GWAS meta-analysis (**Methods**), signals colocalising with genes (*P-*value<5×10^−6^) are plotted for each tissue. **(b) Credible set analysis of RG associations in the European meta-analysis.** Variants from each of the RG signal credible sets are grouped based on their posterior probability (the percentiles labelled on the sides of the bar). SNP variants with posterior probability >80%, along with their locus names are provided. All variants from the credible set of the primary signals are highlighted in bold.

**Table 1.**
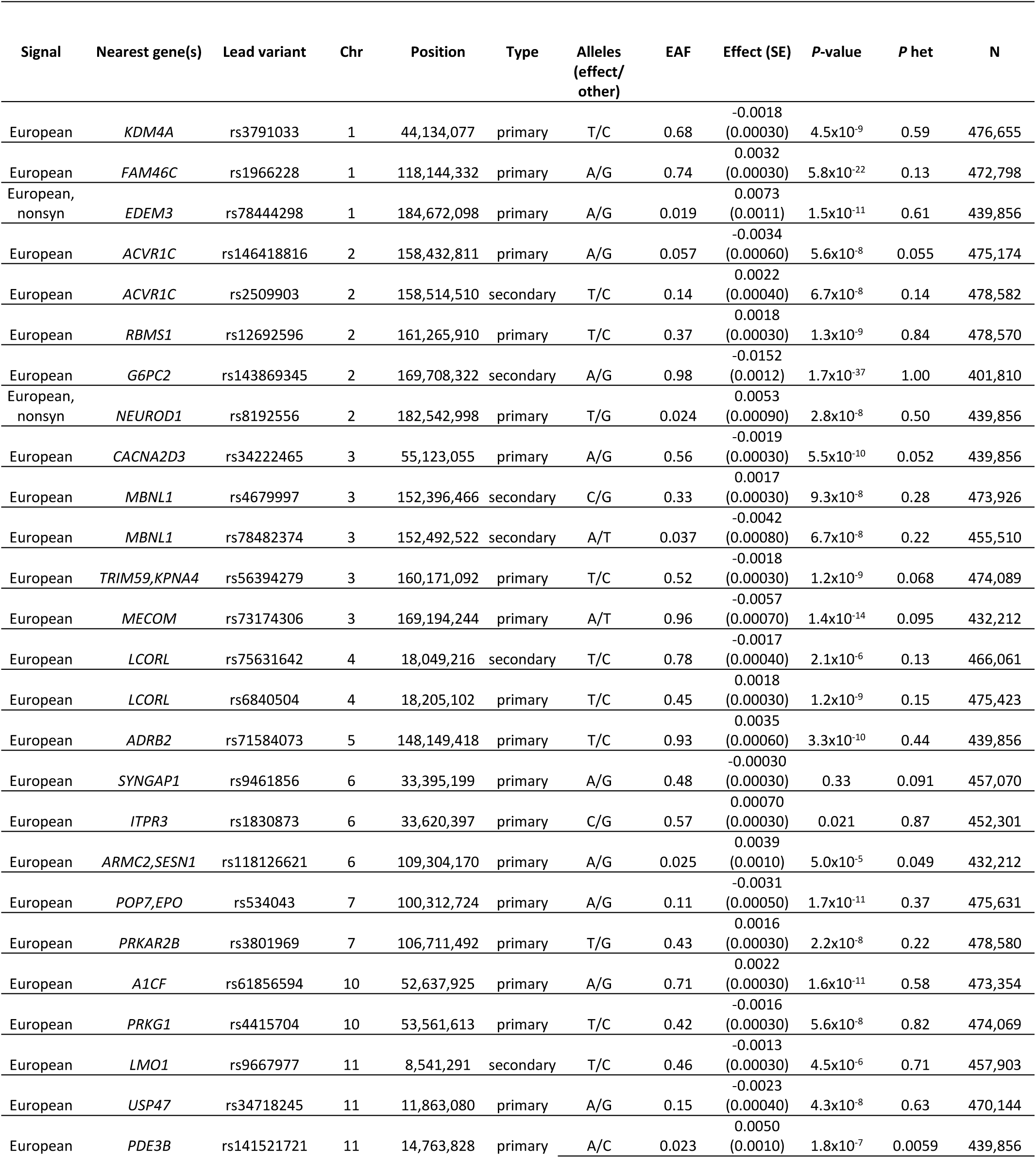
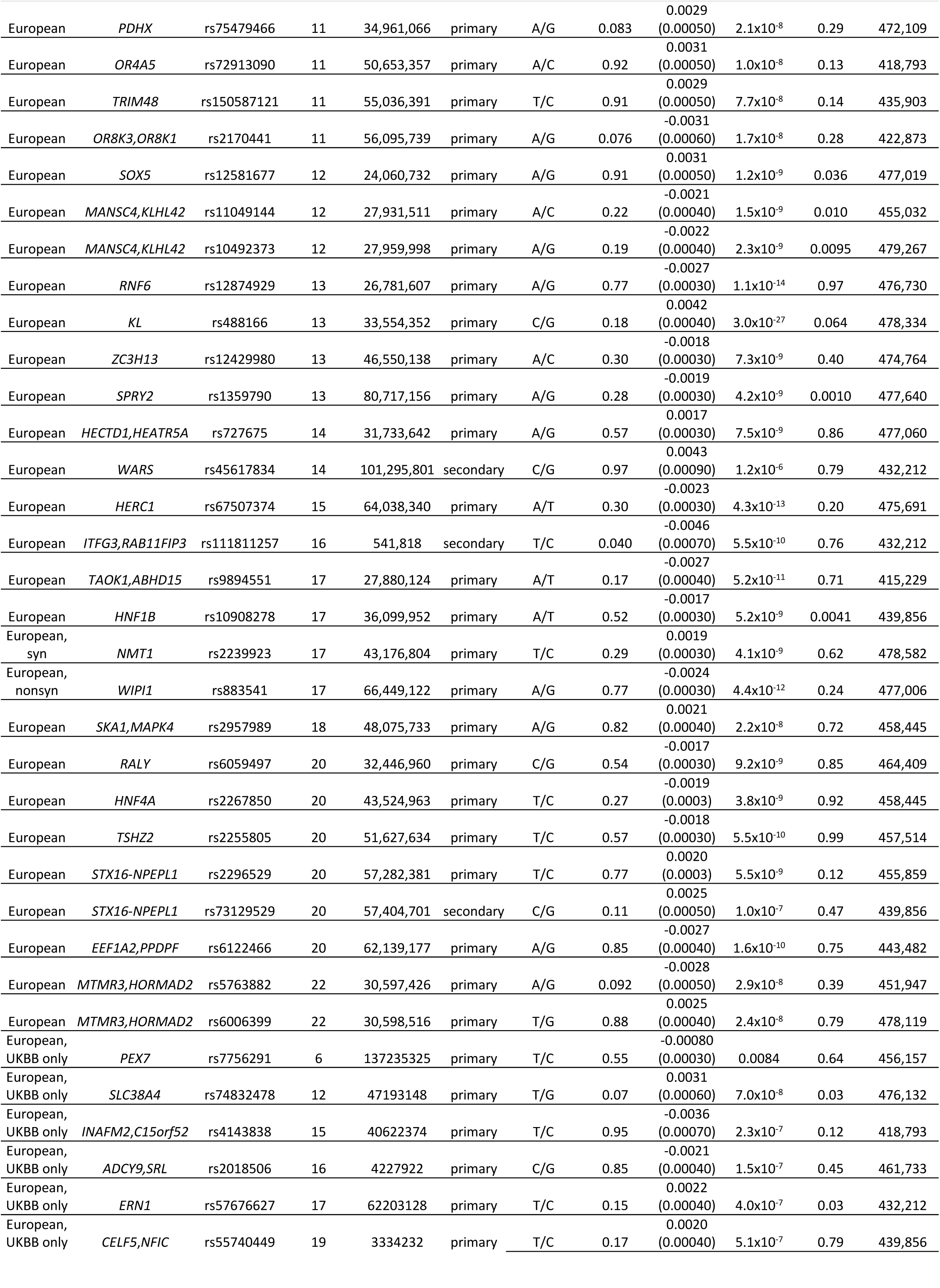
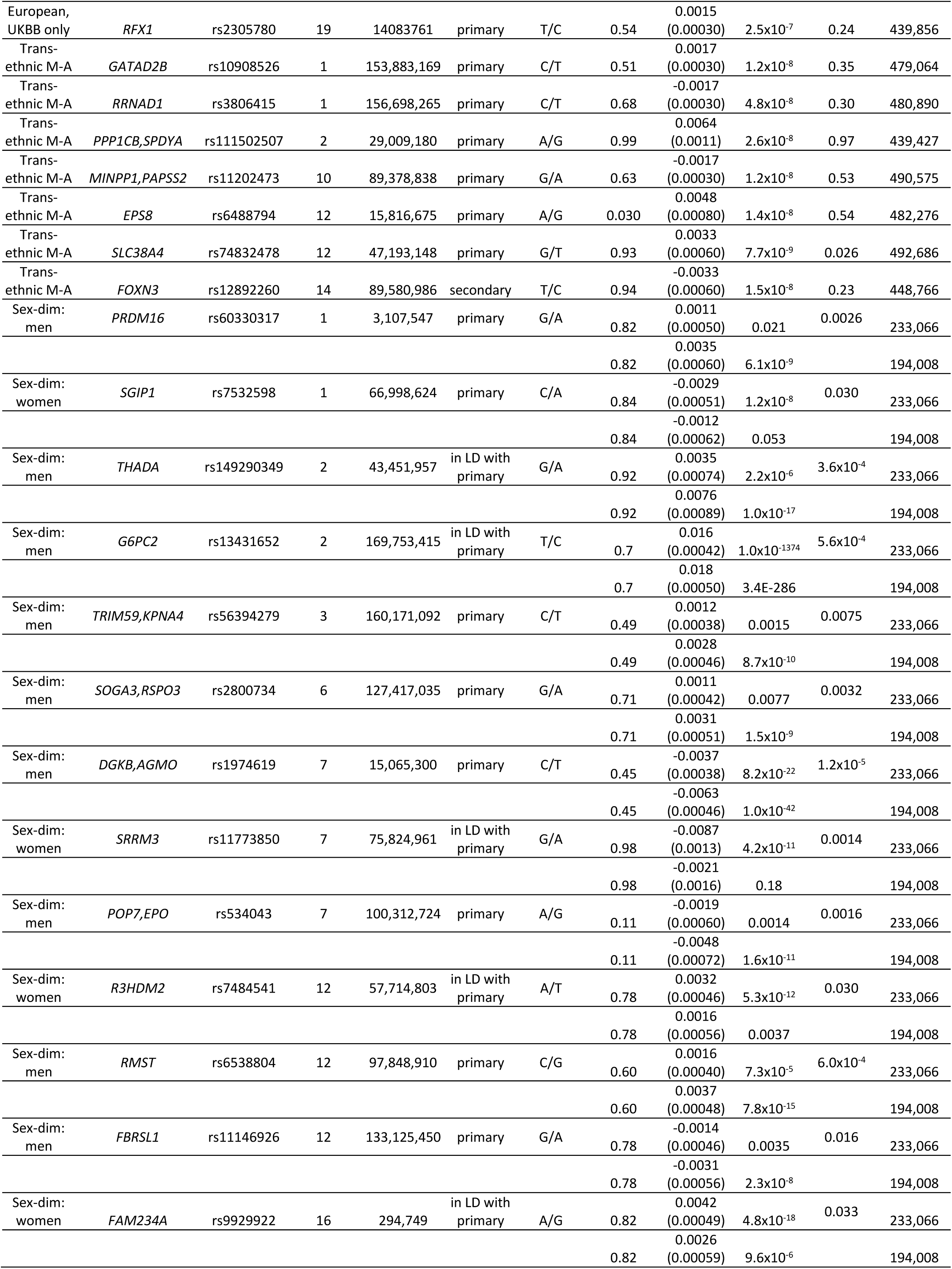
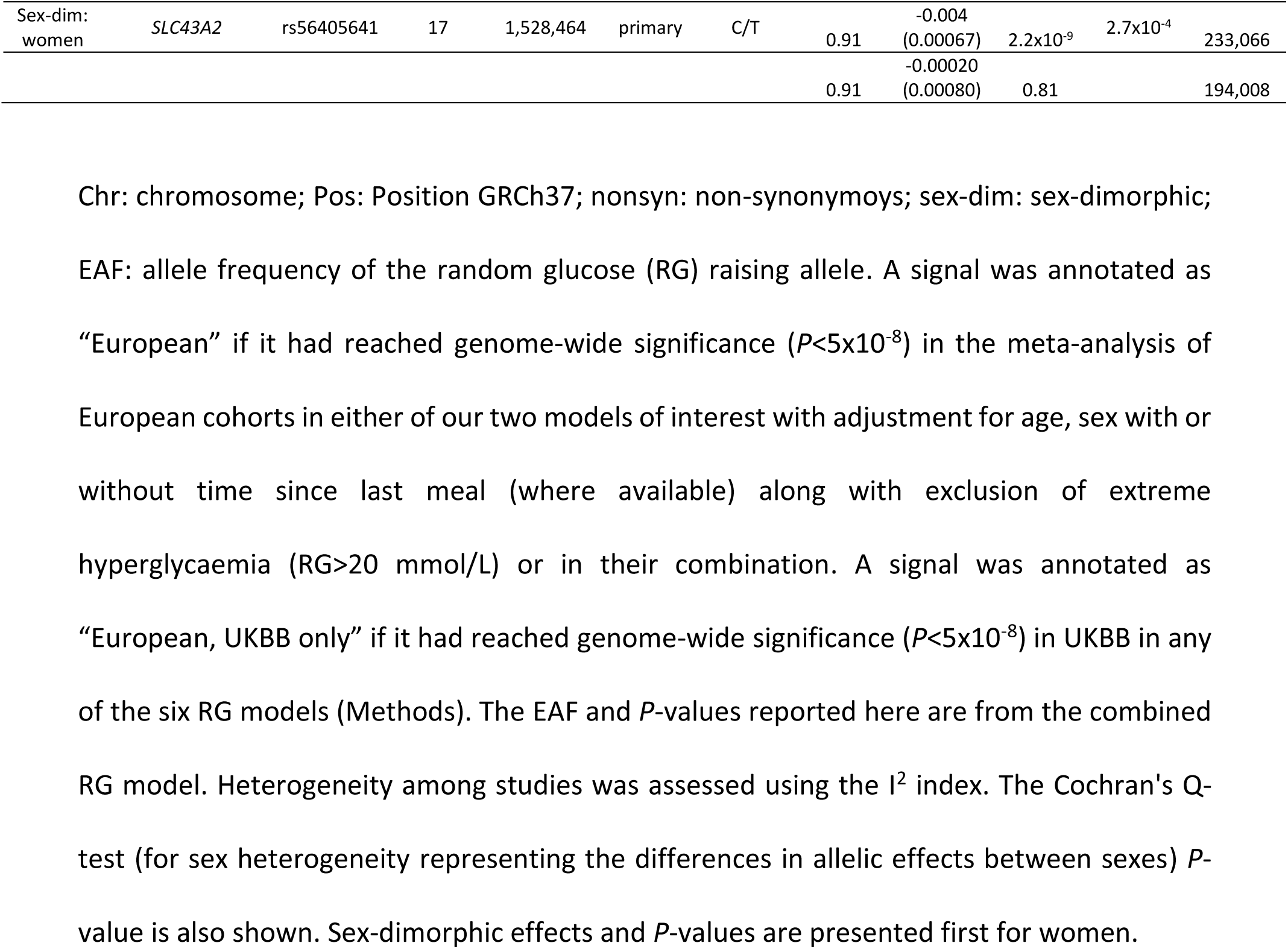
Novel loci for glycaemic traits discovered through i) a GWAS meta-analysis of RG levels in up to 479,482 Europeans without diabetes, and ii) a trans-ethnic meta-analysis of up to 496,036 Europeans and individuals of other ancestries (Black, Indian, Pakistani, Chinese) in UKBB. Loci showing sex-dimorphic effects on glycaemic trait levels for the first time are also shown.

Several of the 162 signals identified in Europeans showed nominal significance (*P*<0.05) in specific UKBB ethnic groups, with *GCK* (rs2908286, r^2^_1000GenomesAllEthnicities_=0.83 with rs2971670 lead in Europeans) reaching genome-wide significance in the African descent individuals alone (**Supplementary Table 3**). Among the novel RG signals, *USP47* was nominally significant in the individuals of African, *FAM46* and *ACVR1C* in the Indian and *TRIM59/KPNA4* and *ZC3H13* in Chinese UKBB ancestry. Trans-ethnic meta-analyses combining Europeans and the other four UKBB ancestral groups revealed seven novel RG signals, including those at *FOXN3, EPS8* and *ISG20L2* (**Table 1**). Overall, while being only 16,554 individuals larger in sample size than the European meta-analysis, the trans-ethnic analysis expanded the novel locus discovery for RG by one tenth (**Supplementary Table 5**).

Among established glycaemic trait signals, the well-known FG loci *G6PC2* (*P*<5.86×10^−754^) and *GCK* (*P*<6.93×10^−301^), with key roles in gluconeogenesis^12^ and glucose sensing^13^, respectively, showed the strongest associations with RG (**Supplementary Table 3**). We also observed two thirds of RG signals overlapping with T2D-risk loci (**Supplementary Figure 1e**), including *SLC30A8, DGKB, TCF7L2, GRB10* and *THADA*. The direction of effects at these loci between RG, T2D and homeostasis model assessment of beta-cell function/insulin resistance (HOMA-B/-IR)^6^ (**Supplementary Figures 1e-f and 2**, **Supplementary Table 6**) were consistent with their epidemiological correlation. Notably, 14 established^14, 15^, such as *DGKB*, *THADA*, *RSPO3*, *G6PC2,* and novel, including *TRIM59*, *POP7*, *SLC43A2*, and *SGIP1*, loci showed sex-dimorphic effects (**Methods, Table 1**, **Figure 1a**, **Supplementary Table 3**). Fine-mapping the associations at RG loci through conditional analysis (**Table 1**) we found three independent coding nonsynonymous rare (minor allele frequency, MAF<1%) variants at *G6PC2* with predicted (rs2232326) and established (rs138726309, rs2232323)^16^ deleterious effects (**Supplementary Table 7**). Within *GCK*, we observed five rare independent (r^2^ <0.001) non-deleterious variants associated with RG at genome-wide significance, including a novel 3’UTR rs2908276 for T2D, glycaemic traits or obesity (**Supplementary Table 7**).

**Figure 2.**
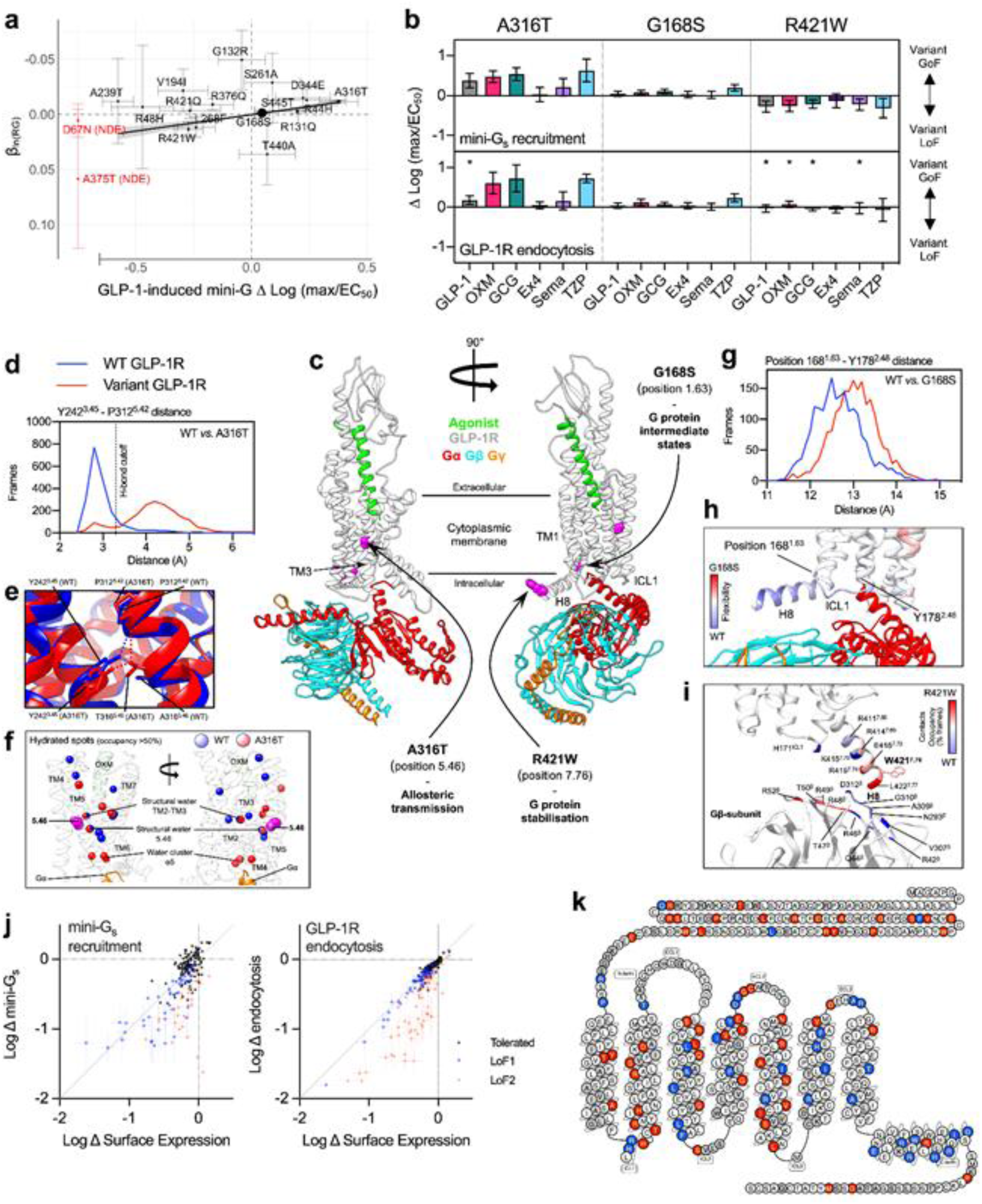
Functional and structural analysis of coding *GLP1R* variants. (**a**) Weighted regression of AST20 β_RG_ estimated in the UKBB study on *GLP1R* variant mini-G_s_ response to GLP-1 stimulation, with correction for variant surface expression, *n*=5-13. Size of dots is proportional to the weight (minor allele frequency) in the regression model (**Methods**). Error bars represent standard errors for β_RG_ and mini-G_s_ coupling in response to GLP-1 stimulation. The grey shaded area corresponds to the 95% confidence interval of the slope of the regression analysis (β=−0.027, 95%CI[-0.036 –{−0.016}], *P*-value=0.0001), which explained 65% of the variance in these associations. Variants in red showed no detectable surface expression (NDE) and are not included in regression analysis. (**b**) *GLP1R* variant mini-G_s_ coupling and receptor endocytosis, with surface expression correction, in response to GLP-1, oxyntomodulin (OXM), glucagon (GCG), exendin-4 (Ex4), semaglutide (Sema) and tirzepatide (TZP), *n*=6. Positive deviation indicates variant gain-of-function, with statistical significance inferred when the 95% confidence intervals shown do not cross zero. Responses are also compared between pathways by unpaired t-test, with * indicating statistically significant differences. (**c**) Architecture of the complex formed between the agonist-bound GLP-1R and Gs; the likely effect triggered by residues involved in GLP-1R isoforms A316T, G168S, and R421W (in magenta) are reported. (**d**) Distributions of the distance between Y242^3.45^ side chain and P312^5.42^ backbone computed during MD simulations of GLP-1R WT and A316T; the cut-off distance for hydrogen bond is shown. (**e**) Difference in the hydrogen bond network between GLP1-R WT and A316T. (**f**) Analysis of water molecules within the TMD of GLP1-R WT and A316T suggests minor changes in the local hydration of position 5.46 (unperturbed structural water molecule). (**g**) Distributions of the distance between position 168^1.63^ and Y178^2.48^ during molecular dynamics simulations of GLP-1R WT and G168S. (**h**) During MD simulations the GLP-1R isoform S168G showed increased flexibility of ICL1 and H8 compared to WT, suggesting a different influence on G protein intermediate states. (**i**) Contact differences between Gs and GLP-1R WT or W421R; the C terminal of W421R H8 made more interactions with N terminal segment of Gs β subunit. (**j**) Mini-G_s_ and GLP-1R endocytosis responses to 20 nM exendin-4, plotted against surface GLP-1R expression, from 196 missense *GLP1R* variants transiently transfected in HEK293T cells (*n*=5 repeats per assay), with data represented as mean ± standard error after normalization to wild-type response and log_10_-transformation. Variants are categorised as “LoF1” when the response 95% confidence interval falls below zero or “LoF2” where expression-normalised 95% confidence interval falls below zero. (**k**) GLP-1R snake plot created using gpcr.com summarizing the functional impact of missense variants; for residues with >1 variant, classification is applied as LoF2>LoF1>tolerated.

Next, we sought to pinpoint the most plausible set of causal variants by calculating 99% credible sets for each of RG loci. In the Europeans only analysis, 19 RG signals were explained by one variant with posterior probability of ≥99% of being causal. For another 20 signals, a lead variant had a posterior probability >80% (**Figure 1b, Supplementary Table 8**). The credible sets were narrowed down in trans-ethnic RG meta-analysis (median credible set size 12.5 in the Europeans only, and 11.0 in the trans-ethnic analysis) (**Supplementary Tables 9 and 10**). This analysis helped to prioritise *GLP1R* for functional studies, in addition to the already deeply characterised *G6PC2* and *CCND2*^17^, all three with lead SNPs of low frequency (1%≤MAF<5%) and posterior probability >99% of being causal.

The lead RG-associated SNPs at *GLP1R, NEUROD1,* and *EDEM3* loci in our analysis were low-frequency coding variants (**Supplementary Figure 3**). *NEUROD1* (Neuronal Differentiation 1) and *EDEM3* (ER Degradation Enhancing Alpha-Mannosidase Like Protein 3) are plausible candidates for glucose homeostasis with the former reported for glucosuria^18^ and the latter linked to renal function^19, 20^. Additionally, lead variants at three previously reported for FG (*GCKR, TET2* and *RREB1*) and two novel RG (*NMT1*, *WIPI1*) loci were all common (MAF≥5%) coding variants (**Supplementary Figure 3**).

### Functional and structural characterisation of RG-associated *GLP1R* coding variants provides a possible framework for T2D treatment stratification

The *GLP1R* gene, identified in our analysis and in previous T2D^21^ and glycaemic trait^22^ GWAS, encodes a class B G protein-coupled receptor (glucagon-like peptide-1 receptor; GLP-1R) that is an established target for glucose-lowering and weight loss in T2D using drugs such as exenatide (exendin-4) and semaglutide^23^. Within *GLP1R,* the lead missense variant at rs10305492 (A316T) had a strong (0.058 mmol/l per allele) RG-lowering effect, second by size only to *G6PC2* locus variants. Previous attempts to functionally characterise A316T and further *GLP1R* variants experimentally have been inconclusive^24^, so we adopted a strategy based on measuring ligand-induced coupling to mini-Gα_s_, representing the most proximal part of the Gα_s_-adenylate cyclase-cyclic adenosine monophosphate (cAMP) pathway that links GLP-1R activation to insulin secretion. Mini-Gα_s_ coupling efficiency was predictive of RG effect for 16 *GLP1R* coding variants detected in the UKBB dataset with effect allele frequency ranging from common (G168S, rs6923761, *P*=4.40×10^−5^) to rare (R421W, rs146868158, *P*=0.054) (**Figure 2a, Supplementary Table 11**), thereby linking differences in experimentally measured GLP-1R function to blood glucose homeostasis.

To probe whether *GLP1R* coding variation could be therapeutically as well as physiologically relevant, we also measured responses to several endogenous and pharmacological GLP-1R agonists. Focussing on the two directly genotyped *GLP1R* missense variants in UKBB, we observed that A316T (rs10305492-A) showed increased responses, and R421W (rs146868158-T) showed reduced responses, to all ligands except exendin-4 (both variants) and semaglutide (A316T only), in line with their RG effects (**Figure 2b**). Agonist-induced GLP-1R endocytosis with R421W was normal despite its signalling deficit, suggestive of biased agonism^26^. The imputed common G168S variant, with relatively small RG-lowering effect (β=−0.0013 [SE=3.14×10^−4^]), also showed subtle increases in function.

To gain structural insights into *GLP1R* variant effects we performed molecular dynamics simulations of the human GLP-1R bound to oxyntomodulin^27^ (**Extended Data Tables 1-6**). A316T has a single amino acid substitution in the core of the receptor transmembrane domain (**Figure 2c**) that leads to an alteration of the hydrogen bond network in close proximity (**Video S1**). In A316T, residue T316^5.46^ replaced Y242^3.45^ in a persistent hydrogen bond with the backbone of P312^5.42^ one turn of the helix above T316^5.46^ (**Figures 2d-e, Video S1**). This triggers a local structural rearrangement that could transmit to the intracellular G protein binding site through transmembrane helix 3 (TM3) and TM5. A structural water molecule was found close to position 5.46 in both A316T and WT (water cluster α5, **Figure 2f**). The same water bridged the backbone of Y241^3.44^ and A316^5.46^ in WT, or the backbone of Y241^3.44^ and the side chain of T316^5.46^ in A316T. Given the importance of conserved water networks in the process of activation of class A GPCRs^28, 29^, the presence of a stable hydrated spot close to position 5.46^30^ corroborates this site as important for tuning the intracellular conformational landscape of GLP-1R. Also, a stabilising role for the water molecules at the binding site of the G protein (water cluster apha5, **Figure 2f**) cannot be ruled out. Note that our results differ from a previous analysis of A316T dynamics^22^, which used an early model that does not fully capture the full structural features of the current active GLP-1R models.

In analogy with A316T, molecular simulations with the G168S variant indicate the formation of a stable new hydrogen bond between the side chain of residue S168^1.63^ and A164^1.59^, located one turn above on the same helix (**Video S2,** **Figure 2g**). This moved the C-terminal end of TM1 closer to TM2 and reduced the overall flexibility of ICL1 (**Figure 2h**), which could potentially alter the role of ICL1 in G protein activation. In contrast to A316T and G168S, the site of mutation R421W is consistent with persistent interactions with the G protein. Simulations predicted a propensity of R421W to interact with a different region of the G protein β-subunit to that engaged by WT (**Figure 2i**).

For a broader view of the impact of *GLP1R* coding variation, we screened an additional 178 missense variants identified from exome sequencing^31^ for exendin-4-induced mini-G_s_ coupling and endocytosis (**Figures 2j-k, Supplementary Table 12**). 110 variants showed a reduced response in either or both pathways (“LoF1”), and 67 displayed a specific response deficit that was not fully explained by differences in GLP-1R surface expression (“LoF2”), with many of these defects being larger than in the analysis in **Figure 2a**.

Overall, these data suggest *GLP1R* variation influences blood glucose levels in health and is likely to be a direct modifier of responses to drug treatment^32^. As some patients fail to respond adequately to GLP-1R agonist treatment, and others are particularly sensitive to side effects^33^, this approach may feed into optimised treatment selection in T2D.

### Functional annotation of RG associations and intestinal health

Previous T2D and glycaemic trait GWAS have primarily implicated pancreatic, adipose and liver tissues^3^. To leverage our RG GWA results to identify additional cell and tissue types with aetiological roles in glucose metabolism, we performed a range of complementary functional annotation analyses in relation to RG GWAS. DEPICT^34^, which predicts enriched tissue types from prioritised gene sets (**Methods**), highlighted intestinal tissues including ileum and colon, as well as pancreas, adrenal glands, adrenal cortex and cartilage (False Discovery Rate<0.20) (**Figures 3a-b, Supplementary Tables 13a-c**). Similarly, CELLECT^35^, which facilitates cell-type prioritisation based on single cell RNAseq datasets (**Methods**), identified large intestinal tissue as the second ranked only to pancreatic cell types (**Figure 3c, Supplementary Table 14)**; interestingly, RG variants were related particularly to enriched expression in pancreatic polypeptide (PP) cells, exceeding even the more conventionally implicated insulin-secreting beta cells. Supporting evidence was obtained from transcriptome-wide association study (TWAS) analysis (**Methods**), where we identified a total of 216 (119 unique) significant genetically driven associations across the ten tested tissues; 52 (26 unique) of highlighted genes are located at genome-wide significant RG loci (**Supplementary Tables 15a)**. TWAS signals in skeletal muscle showed the largest overlap with RG signals, such as *GPSM1*^36^ and *WARS*; with combined results from ileum and colon also highly enriched, including the novel *NMT1* and the established *FADS1/3* and *MADD* genes (**Figure 1a, Supplementary Tables 15a-b**). Moreover, epigenetic annotations using the GARFIELD tool highlighted significant (*P*<2.5×10^−5^, **Methods**) enrichment of RG-associated variants in foetal large intestine, as well as blood, liver and other tissues (**Supplementary Figure 4, Supplementary Table 16**). Adult intestinal tissues are not available in GARFIELD except for colon. Prompted by multiple analyses highlighting a potential role for the digestive tract in glucose regulation, we assessed the overlap between our signals and those from the latest microbiome GWAS^37^ (**Methods**) and identified three genera sharing signals with RG at two loci: *Collinsella* and *LachnospiraceaeFCS020* at *ABO-FUT2* and *Slackia* at *G6PC2* (**Figure 1a, Supplementary Table 17**). The *ABO-FUT2* locus effects on RG could be mediated by abundance of bacteria producing glucose from lactose and galactose^38^.

**Figure 3.**
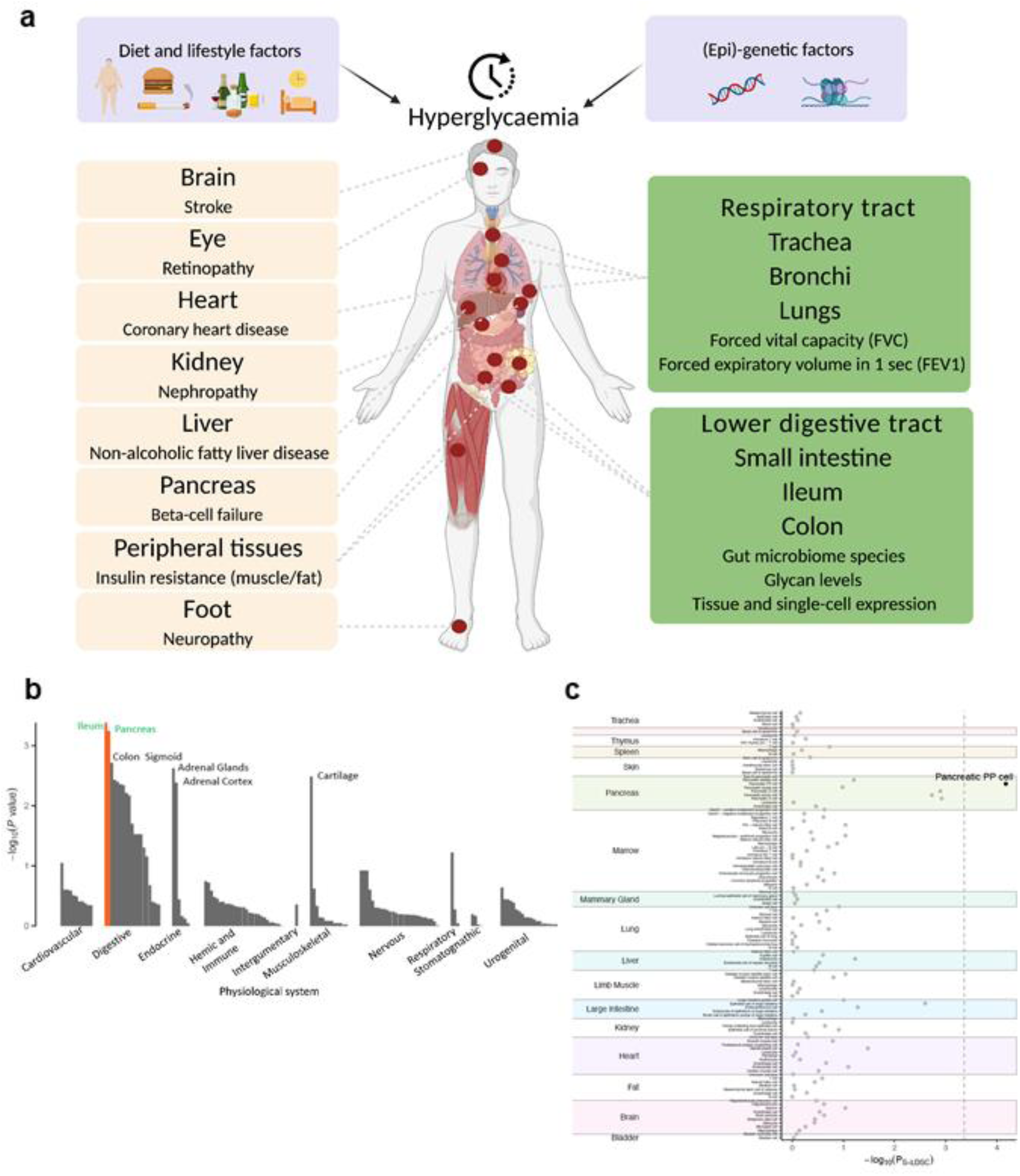
Deterioration of glucose homeostasis progressing into type 2 diabetes (T2D) and leading to complications in multiple organs and tissues - established (left, in peach colour) and new (right, in green). (**a**) A human figure illustrating the main causes of hyperglycemia (a combination of lifestyle and genetic factors), and how hyperglycemia affects many organs and tissues. Complications on the left panel are well established for T2D. Those on the right panel are emerging ones and are supported by our current analyses. (**b**) Functional annotation of the RG GWAS results with DEPICT (**Methods**). (**c**) Functional annotation of the RG GWAS results with CELLECT (**Methods**).

eQTL colocalization analyses, using eQTLgen blood expression data from 31,684 individuals^39^ and the COLOC2 approach (**Methods**), identified 14 loci with strong links (posterior probability >50%) to gene expression data, including *SMC4*, *TRIM59*, *EIF5A2*, *TET2*, *COG5*, *CHMP5*, *NFX1*, *FNBP4*, *MADD*, *RAPSN*, *WARS1*, *HBM*, *NUFIP2*, and *PPDPF* (**Supplementary Table 18**). This further supported elucidation of biological candidates at novel and established glycaemic loci.

Finally, we observed associations at two RG loci (*GCKR, HNF1A*) with nine total plasma N-glycome traits^40^ at a Bonferroni corrected threshold (**Methods,** **Figure 1a, Supplementary Table 19**). These traits represent highly branched galactosylated sialylated glycans (attached to alpha1-acid protein - an acute-phase protein^41^), known to lead to chronic low-grade inflammation^42, 43^ and an increased risk of T2D^44–46^ that might be explained by the role of N-glycan branching of the glucagon receptor in the glucose homeostasis^47^. In addition, ten glycans showed association with five RG loci (*GCKR, HNF1A*, *BAG1*, *PLUT*, *ACVR1C*) loci at a suggestive level of significance (**Figure 1a**). Among them, three are attached to immunoglobulin G molecules^41^ and their increased relative abundances are associated with a lower risk of T2D^48^ and diminished inflammation status^49^.

### Analysis of genetic relationships between RG and other metabolic or non-metabolic traits

To quantify the shared genetic contribution between RG and other phenotypes, we estimated their genetic correlations using linkage-disequilibrium score regression analyses. We detected positive genetic correlations between RG, squamous cell lung cancer (r_g_=0.28, *P*=0.0015), and lung cancer (r_g_=0.12, *P*=0.037, **Figure 4, Supplementary Table 20**); as well as inverse genetic correlations with lung function related traits, such as forced vital capacity (FVC, r_g_=−0.090, *P*=0.0059) and forced expiratory volume in 1 second (FEV1, r_g_=−0.054, *P*=0.017) (**Figures 3a and 4, Supplementary Table 20**). To investigate this further, we conducted a bi-directional Mendelian Randomisation (MR) analysis, which suggested a causal effect of RG and T2D on lung function, including FEV1 (β_MR-RG_=−0.60, *P*=0.0015; β_MR-T2D_=−0.049, *P*=1.27×10^−13^) and FVC (β_MR-RG_=−0.61, *P*=3.5×10^−4^; β_MR-T2D_=−0.062, *P*=1.42×10^−21^), but not *vice versa* (**Methods, Supplementary Table 21**). Previous observational studies have highlighted worsening lung function, as defined by FVC, in T2D patients^50, 51^. More recently, it was shown that patients with diabetes are at an increased risk of death from the viral infection COVID-19^52^, with pulmonary dysfunction contributing to mortality^53^. Our data therefore support the causal effect of glycaemic dysregulation on a decline in lung function as a novel complication of diabetes.

**Figure 4.**
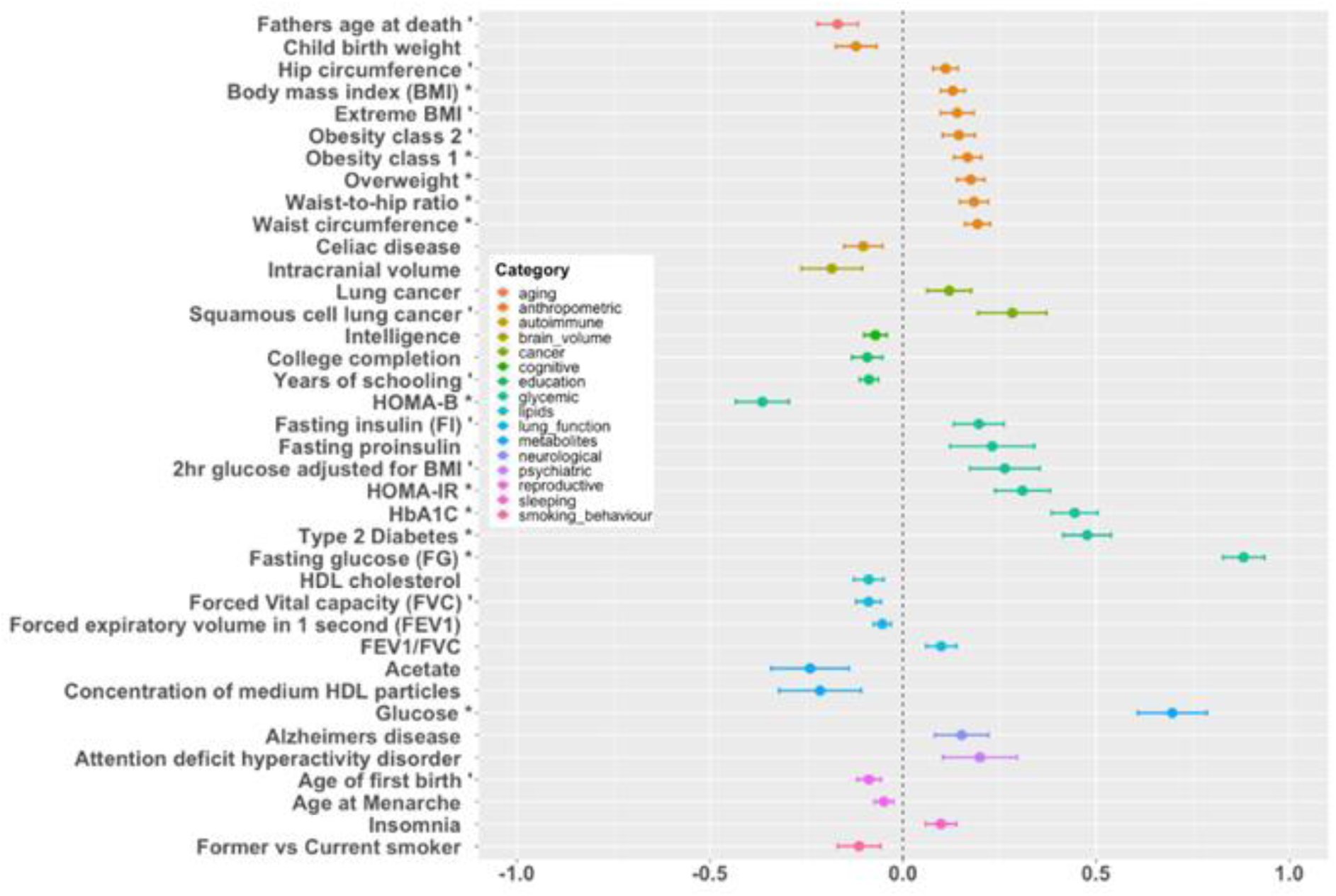
Genome-wide genetic correlation between RG and a range of traits and diseases. X axis provides the r_g_ genetic correlation values for traits or diseases (Y axis) reaching at least nominal significance. Correlations reaching a *P-*value<0.01 are labelled with “ ‘ “, and those *P-*value<0.05/239 are labelled with “ * “.

Genome-wide genetic correlation analyses also showed strong positive genetic correlation of RG with FG (r_g_=0.88, *P*=6.93×10^−61^, **Figure 4, Supplementary Table 20**). We meta-analysed RG studies other than UKBB with FG GWAS summary statistics^54^, observing 77 signals reaching nominal significance that were directionally consistent in both UKBB and RG+FG (**Supplementary Table 3**), providing an additional support to our RG findings. Given the large genetic overlap between RG, other glycaemic traits and T2D, we evaluated the ability of a trait-specific polygenic risk score (PRS) to predict RG, T2D and glycated haemoglobin (HbA1c) levels using UKBB effect estimates and the Vanderbilt cohort (**Methods**). The RG PRS explained 0.58% of the variance in RG levels when individuals with T2D were included, (**Supplementary Table 22**) and 0.71% of the variance after excluding those who developed T2D within one year of their last RG measurement. The RG PRS performance was comparable to that of the FG loci PRS (0.38% vs. 0.42% for T2D; 0.40% vs. 0.44% for HbA1c) indicating wide similarities with the latter.

We previously highlighted diverse effects of FG and T2D loci on pathophysiological processes related to T2D development by grouping associated loci in relation to their effects on multiple phenotypes^6^. Cluster analysis of the RG signals with 45 related phenotypes identified three separate clusters that give insights into the aetiology of glucose regulation and associated disease states (**Methods,** **Figure 1a, Supplementary Table 23, Supplementary Figures 5a-d**). Cluster 1 (“metabolic syndrome” cluster) clearly separated 33 loci with effects on higher waist-to-hip ratio, blood pressure, plasma triglycerides, insulin resistance (HOMA-IR) and coronary artery disease risk, as well as lower testosterone and sex hormone binding globulin levels in men. Cluster 3 was characterised in particular by insulin secretory defects^6^. Cluster 2 was less clearly defined by a primary effect on insulin release *versus* insulin action^3^, but interestingly included a sub-cluster of 21 loci which exert protective effects on inflammatory bowel disease. Moreover, cluster 2 was notable for generally reduced impact on T2D risk in comparison to clusters 1 and 3, underscoring the partial overlap between genetic determinants of glycaemia and T2D that is known to exist^55^.

### Discussion

Taking advantage of data from 493,036 individuals, we have expanded by 58 the number of loci associated with glycaemic traits. By using RG, our analysis integrates genetic contributions to a wider range of physiological stages than possible with FG or other standardised measures. Moreover, the greater statistical power obtained from large trans-ethnic meta-analysis improves confidence in identification of potentially causal variants, thereby helping to prioritise loci for more detailed functional analyses in the future. Our observation of ligand-specific responses to the A316T, G168S and R421W *GLP1R* variants provides a mechanism that can explain why some individuals respond better or worse to particular GLP-1R-targeting drugs. We note that other class B GPCRs identified in our current analysis and other glycaemic or T2D GWAS include *GIPR*, *GLP2R*^3^ and *SCTR*^21^, all of which are investigational targets for T2D treatment. Our functional annotation analyses point to underexplored tissue mediators of glycaemic regulation, with several sources of evidence highlighting a likely role of the intestine. This observation is compatible with the well-described and profound effects of gastric bypass surgery on T2D resolution^56^, as well as links between the intestinal microbiome and responses to several diabetes drugs^57^. Finally, through Mendelian randomisation we were able to identify a causal effect of glucose levels and T2D on lung function, demonstrating the utility of this approach for the corroboration of findings from observational studies and elevating lung dysfunction as a new complication of diabetes.

### Methods

#### Phenotype definition and model selection for RG GWAS

We used RG (mmol/l) measured in plasma or in whole blood (corrected to plasma level using the correction factor of 1.13). Individuals were excluded from the analysis, if they had a diagnosis of T2D or were on diabetes treatment (oral or insulin). Individual studies applied further sample exclusions, including pregnancy, fasting plasma glucose equal to or greater than 7 mmol/l in a separate visit, when available, and having type 1 diabetes. Detailed descriptions of study-specific RG measurements are given in **Supplementary Table 1**. All studies were approved by local ethics committees and all participants gave informed consent. We examined the distributions of untransformed and natural logarithmic transformed RG in the first set of six available cohorts. We observed that RG was approximately normally distributed after natural log transformation. We then determined the variables that could have a significant effect on RG by fitting several regression models using naturally log-transformed RG as the outcome with age, sex, BMI and time since last meal as predictors. Modelling of RG revealed significant effects (*P*<0.05) of age, sex, BMI and time since last meal (accounted for as T, T^2^ and T^3^) in these cohorts (**Supplementary Table 2**). Compared to RG models without T, inclusion of T, T^2^ and T^3^ increased the proportion of variance explained in the range of 1-6%. Thus, inclusion of this covariate is potentially equivalent to 1-6% increase in study sample size. For the GWAS, we included individuals based on two RG cut-offs: <20 mmol/l (20) to account for the effect of extreme RG values and <11.1 mmol/l (11), which is an established threshold for T2D diagnosis. We then evaluated six different models in GWAS according to covariates included and cut-offs used: 1) age (A) and sex (S), RG<20 mmol/L (**AS20**), 2) age, sex and BMI (B), RG<20 mmol/L (**ASB20**), 3) age and sex, RG<11.1 mmol/L (**AS11**), 4) age, sex and BMI, RG<11.1 mmol/L (**ASB11**), 5) age, sex, T, T^2^ and T^3^, RG<20 mmol/L (**AST20**) and 6) age, sex, T, T^2^ and T^3^ and BMI, RG<20 mmol/L (**ASTB20**). Apart from above, additional adjustments for study site and geographical covariates were also applied.

#### Genotyping and quality control

Commercial genome-wide arrays and the Metabochip^58^ were used by individual studies for genotyping. Studies with genome-wide arrays undertook imputation of missing genotypes using at least the HapMap II CEU reference panel via MACH^59^, IMPUTE^60^ or MINIMAC^61^ software (**Supplementary Table 1**). For each study, samples reflecting duplicates, low call rate, gender mismatch, or population outliers were removed. Low-quality SNPs were excluded by the following criteria: call rate <0.95, minor allele frequency (MAF) <0.01, minor allele count <10, Hardy-Weinberg *P*-value <10^−4^. GWAS were performed with PLINK, SNPTEST, EMMAX, R package LMEKIN, Merlin, STATA, and ProbABEL (**Supplementary Table 1**).

#### GWAS in the UKBB

For the GWAS of the UKBB data we excluded non-white non-European individuals and those with discrepancies in genotyped and reported sex. For the RG definition, we used the same criteria as in the other studies described above. To control for population structure, we adjusted the analyses for six first principal components. The GWAS was performed using the BOLT-LMM v2.3 software^62, 63^ restricting the analyses to variants with MAF>1% and imputation quality>0.4.

#### RG meta-analyses

The GWAS meta-analysis of RG consisted of four components: (i) 37,239 individuals from 10 European GWAS imputed up to the HapMap 2 reference panel, (ii) 3,156 individuals from three European GWAS with Metabochip coverage, (iii) 21,083 individuals from two European GWAS imputed up to 1000 genomes reference panel and iv) 401,810 individuals of white European origin from the UKBB and (iv) 16,983 individuals from the Vanderbilt cohort imputed to the HRC panel. We imputed the GWAS meta-analysis summary statistics of each component to all-ancestries 1000 Genomes reference panel^64^ using summary statistics imputation method implemented in the SS-Imp v0.5.5 software^65^. SNPs with imputation quality score <0.7 were excluded. We then conducted inverse variance meta-analyses to combine the association summary statistics from all components using METAL (version from 2011-03-25)^66^. We focused our meta-analyses on models AS20 (17 cohorts, N_max_=481,150) and AST20 (when time from last meal was available in the cohort) (12 cohorts, N_max_=438,678). For FHS cohort, where no information was available for individuals with RG>11.1 (an established threshold for 2hGlu concentration, which is a criterion for T2D diagnosis), AS11 model results were used. In order to maximise the association power while taking into account T, we also performed meta-analysis using AST20 (when time from last meal was available in the cohort) combined with AS20 (otherwise) and we termed this analysis as AS20+AST20 in the following text (17 cohorts, N_max_=480,250).

A signal was considered to be associated with RG if it had reached genome-wide significance (*P*<5×10^−8^) in the meta-analysis of UKBB and other cohorts in either of our two models of interest (AS20) or (AST20) or in their combination (AS20+AST20). We report the *P*-value from the combined model, unless otherwise stated. Full results from all models are provided in the **Supplementary Table 3**. All the follow-up analyses were conducted using the combined AS20+AST20 model. We checked for nominal significance (*P*<0.05) and directional consistency of the effect sizes for the selected leads in the combined model in UKBB results vs other cohort results. We further extended the check between UKBB results and meta-analysis of other cohorts including FG GWAS meta-analysis^54^ excluding overlapping cohorts. This meta-analysis conducted in METAL was sample size and *P*-value based due to the measures being at different scale (natural logarithm transformed RG and untransformed FG).

#### Trans-ethnic analyses and meta-analysis

We performed GWAS in those non-European populations within UKBB that had a sample size of at least 1,500 individuals. These were Black (N=7,644), Indian (N=5,660), Pakistani (N=1,747) and Chinese (N=1,503). We further meta-analysed our European cohorts with the trans-ethnic UKBB cohorts. The analyses were performed with BOLT-LMM and METAL.

#### Sex-dimorphic analysis

To evaluate sex-dimorphism in our results, we meta-analysed the UKBB and the Vanderbilt cohort with the GMAMA software^67^, which provides a 2 degrees of freedom (df) test of association assuming different effect sizes between the sexes. We considered a signal to show evidence of sex-dimorphism if the 2 df test *P*-value was <5×10^−8^ and if the sex heterogeneity *P*-value (1 df) was <0.05.

#### Clumping and GCTA analysis

We performed a standard clumping analysis [PLINK 1.9 (v1.90b6.4)^68^ criteria: *P*≤5×10^−8^, r^2^=0.01, window-size=1Mb, 1000 Genomes Phase 3 data as linkage disequilibrium (LD) reference panel] to select a list of near-independent signals. We then performed a stepwise model selection analysis (GCTA conditional analysis) to replicate the analysis using GCTA v1.93.0^69^ with the following parameters: *P*≤5×10^−8^ and window-size=1Mb. We further checked for additional distinct signals by using a region-wide threshold of *P*≤1×10^−5^ for statistical significance.

#### GLP-1R pharmacological and structural analysis

##### Reagents

Custom peptides were purchased from Wuxi Apptec and were at least 95% pure. SNAP-Surface probes were purchased from New England Biolabs. BG-S-S-649^70^ was provided by New England Biolabs on a collaborative basis. Furimazine was obtained from Promega.

##### Plasmids and cell line generation

Wild-type and variant GLP-1R expression plasmids, termed pcDNA5-SNAP_f_-GLP-1R-SmBiT, were generated by Genewiz, as previously described^71^, to the following design: a fast-labelling SNAP_f_ tag and upstream signal peptide based on that of the 5-HT_3A_ receptor (MDSYLLMWGLLTFIMVPGCQA), plus C-terminal SmBiT tag, were appended to the codon-optimised wild-type or variant human GLP-1R sequence (without the endogenous N-terminal signal peptide, which would lead to cleavage of the N-terminal SNAP-tag; accordingly, known missense variants in the signal peptide region were not included), and inserted into the pcDNA5/FRT/TO expression vector. These constructs allow bio-orthogonal labelling of expressed GLP-1R using SNAP-labelling probes and monitoring of cytosolic protein interactions made to GLP-1R. Constructs were used either for transient transfection or to generate stable cell lines. To obtain cell populations with inducible expression of SNAP-GLP-1R-SmBiT from a single genomic locus, Flp-In™ T-REx™ 293 cells^72^ (Thermo Fisher) were co-transfected with pOG44 (Thermo Fisher) and wild-type or variant pcDNA5-SNAP_f_-GLP-1R-SmBiT in a 9:1 ratio, followed by selection with 100 µg/ml hygromycin. The resulting cell lines were maintained in DMEM supplemented with 10% foetal bovine serum (FBS) and 1% penicillin/streptomycin.

#### Mini-G_s_ recruitment assay

Assays were performed as previously described^71^. Where stable cell lines were used (i.e. **Figures 2a and 2b**), wild-type or variant T-REx-SNAP-GLP-1R-SmBiT cells were seeded in 12-well plates and transfected with 1 µg/well LgBiT-mini-G_s_ (a gift from Prof Nevin Lambert, Medical College of Georgia). The following day GLP-1R expression was induced by addition of tetracycline (0.2 µg/ml) to the culture medium for 24 hours. For transient transfection assays (i.e. **Figure 2j**), HEK293T cells in poly-D-lysine-coated white 96-well plates were transfected using Lipofectamine 2000 with 0.05 µg/well wild-type or variant SNAP-GLP-1R-SmBiT plus 0.05 µg/well LgBiT-mini-G_s_ and the assay performed 24 hours later. Cells were then resuspended in Hank’s balanced salt solution (HBSS) + furimazine (Promega) diluted 1:50 and seeded in 96-well half area white plates, or the same reagent added to adherent cells for transient transfection assays. Baseline luminescence was measured over 5 min using a Flexstation 3 plate reader at 37°C before addition of ligand or vehicle. Agonists were applied at a series of concentrations spanning the response range. After agonist addition, luminescent signal was serially recorded over 30 min, and ligand-induced effects were quantified by subtracting individual well baseline. Signals were corrected for differences in cell number as determined by BCA assay.

#### High content imaging-based GLP-1R internalisation assay

The assay was performed as previously described^71^. Where stable cell lines were used (i.e. **Figures 2a and 2b**), wild-type or variant T-REx-SNAP-GLP-1R-SmBiT cells were seeded (10,000/well) in poly-D-lysine-coated black, clear-bottom 96-well plates, in complete medium supplemented with tetracycline (0.2 µg/ml) for 24 hours before the assay. Medium was removed and cells labelled with 0.5 µM BG-S-S-649 (a gift from New England Biolabs) in complete medium for 20 min at 37°C. Agonists were then applied in serum-free medium at the indicated dose for a 30-min stimulation period to induce GLP-1R internalisation. A series of concentrations spanning the response range were used. Cells were then washed with HBSS, followed by a 5-min treatment ± 100 mM sodium 2-mercaptoethanesulfonate (Mesna) in alkaline TNE buffer (pH 8.6) to cleave residual surface BG-S-S-649 without affecting that internalised whilst bound to SNAP-GLP-1R. After re-washing, the plate was imaged using a 0.75 numerical aperture 20x phase contrast objective, with 9 fields-of-view (FOVs) per well acquired for both transmitted phase contrast and epifluorescence. Flat-field correction of epifluorescence images was performed using BaSiC^73^ and cell segmentation was performed using PHANTAST^74^ for the phase contrast image. To determine specific GLP-1R labelling, cell-free background per image was determined from the segmented epifluorescence image and subtracted from the mean fluorescence intensity from the cell-containing regions. Ligand induced effects were determined by subtracting the signal from vehicle-treated cells exposed to Mesna. Responses were normalised to signal from labelled, untreated cells (i.e. total surface labelling) within the same assay. GLP-1R surface expression levels were also obtained from these assays from wells not treated with GLP-1RA or Mesna. For transient transfection assays (i.e. **Figure 2j**), the assay was performed similarly but with the following changes: 1) HEK293T cells in poly-D-lysine-coated black clear-bottom 96-well plates were transfected using Lipofectamine 2000 with 0.1 µg/well wild-type or variant SNAP-GLP-1R-SmBiT and the assay performed 24 hours later; 2) the plate was imaged as above both prior to and after ligand treatment (+subsequent Mesna cleavage); 3) surface labelling quantification was obtained from the pre-treatment read, and total internalised receptor was obtained from the post-treatment read.

#### Analysis of pharmacological data

Technical replicates within the same assay were averaged to give one biological replicate. For concentration-response assays (**Figures 2a and 2b**), ligand-induced responses were analysed by 3-parameter fitting in Prism 8.0 (GraphPad Software). As a composite measure of agonism^75^, log_10_-transformed E_max_/EC_50_ values were obtained for each ligand/variant response. The wild-type response was subtracted from the variant response to give Δlog(max/EC_50_), a measure of gain- or loss-of-function for the variant relative to wild-type. Log_10_-transformed surface expression levels were obtained for each variant relative to wild-type; these were then used to correct mini-G_s_ Δlog(max/EC_50_) values for differences in variant GLP-1R surface expression levels, by subtraction with error propagation. GLP-1R internalisation responses were already normalised to surface expression within each assay. Statistical significance between wild-type and variant responses was inferred if the 95% confidence intervals for Δlog(max/EC_50_) did not cross zero^75^. Changes to the profile of receptor response between mini-G_s_ recruitment and GLP-1R internalisation were inferred if p<0.05 with unpaired t-test analysis, with Holm-Sidak correction for multiple comparisons. For transient transfection assays (**Figure 2j**), responses were normalised to wild-type response and log_10_ transformed to give Log Δ response. Additionally, the impact of differences in surface expression on functional responses was determined by subtracting log-transformed normalised expression level from log-transformed normalised response.

#### Variance explained in RG effects by mini-Gs recruitment at coding *GLP1R* variants

RG (AST20) effects estimated in the UKBB study at 18 independent (r^2^<0.02) coding *GLP1R* variants (**Supplementary Table 10**) were regressed on mini-Gs coupling in response to GLP-1 stimulation (corrected for surface expression) giving more weight to variants with higher minor allele frequency. Adjusted R^2^ is reported as variance explained in RG effects by mini-G_s_ coupling.

#### Computational methods including molecular dynamics simulations

The active state structure of GLP-1R in complex with OXM^27^ and Gs protein was modelled as previously described^30^ and used to simulate the WT GLP-1R and G168S, A316T and R421W. The systems were prepared for molecular dynamics (MD) simulations and equilibrated as reported in^30^. AceMD3^76^ was employed for production runs (four MD replicas of 500 ns each). AquaMMapS analysis^77^ was performed as previously described^30^.

#### Credible set analysis

After selecting the signals with each region based on different M-A results from AS20, AST20 and AS20+AST20 models, we further performed a credible set analysis to obtain a list of potential causal variants for each of the 143 selected signals. Based on the method adopted from^78^ under the assumption that there is one causal variant within each region, we created 99% credible sets. We also calculated credible sets for the trans-ethnic meta-analysis and compared the results between the European only and trans-ethnic meta-analyses.

#### DEPICT analysis

DEPICT uses GWAS summary statistics and computes a prioritization of genes in associated loci, which are used to prioritise tissues via enrichment analysis. DEPICT v1 (rel 194) was used with default settings and RG GWAS summary statistics as input against a genetic background of SNPsnap data^79^ derived from the 1000 Genomes Project Phase 3^80^ in order to prioritise genes. Tissue and cell types enriched for prioritised genes were computed on normalised expression data comprised of 209 tissues and cell types from 37,427 Affymetrix U133 Plus 2.0 Array, as previously described^34^. We used 500 permutations for bias adjustment and 50 replications for false discovery rate estimation in our analysis in order to calculate empirical *P*-values and false discovery rate cutoffs for prioritised tissues.

#### CELLECT analysis

CELL type Expression-specific integration for Complex Traits (CELLECT)^35^ v1.0.0 and Cell type EXpression-specificity (CELLEX)^35^ v1.0.0 are two toolkits for genetic identification of likely etiologic cell types using GWAS summary statistics and single-cell RNA-sequencing (scRNA-seq) data. Tabula muris gene expression data^81^, a scRNA-seq dataset derived from 20 organs from adult male and female mice, was pre-processed as described previously^82^. Briefly, expression values were normalised by using a scaling factor of 10k transcripts. The normalised values were transformed by taking log(x+1), followed by filtering out infrequently expressed genes, and keeping only those mouse transcripts with 1-1 mapping to human genes in Ensembl v.91. This data was supplied to CELLEX to compute a cumulative expression specificity metric (ESμ) of every gene for each Tabula muris cell type by combining four different expression specificity measures^82^. ESμ values were converted to stratified LD-score regression (S-LDSC) annotations using the 1000 Genomes Project SNPs and mapping each SNP to the strongest ESμ value within 100kb. Cell types were prioritised by S-LDSC on the basis of ESμ-derived annotations and GWAS summary statistics from the current RG meta-analysis.

#### Genetically regulated gene expression analysis

We used MetaXcan (S-PrediXcan) v0.6.10^83^ to identify genes whose genetically predicted gene expression levels are associated with RG in a number of tissues. The tested tissues were chosen based on their involvement in glucose metabolism. Those were adipose visceral omentum, adipose subcutaneous, skeletal muscle, liver, pancreas and whole blood. Additionally, we tested ileum, transverse colon, sigmoid colon and adrenal gland, because they were highlighted by DEPICT analysis. The models for the tissues of interest were trained with GTEx Version 7 transcriptome data from European individuals^84^. The tissue transcriptome models and 1000 Genomes^85^ based covariance matrices of the SNPs used within each model were downloaded from PredictDB Data Repository. The association statistics between predicted gene expression and RG were estimated from the effects and their standard errors coming from the AS20+AST20 model. Only statistically significant associations after Bonferroni correction for the number of genes tested across all tissues (*P* ≤ 8.996×10^−7^) were included into the table. Genes, where less than 80% of the SNPs used in the model were found in the GWAS summary statistics, were excluded due to low reliability of association result.

#### GARFIELD analysis

We applied the GARFIELD tool v2^86^ on the RG AS20+AST20 meta-analysis results to assess enrichment of the RG-associated variants within functional and regulatory features. GARFIELD integrates various types of data from a number of publicly available cell lines. Those include genetic annotations, chromatin states, DNaseI hypersensitive sites, transcription factor binding sites, FAIRE-seq elements and histone modifications. We considered enrichment to be statistically significant if the RG GWAS *P*-value reached *P*=1×10^−8^ and the enrichment analysis *P*-value was <2.5×10^−5^ (Bonferroni corrected for 2040 annotations).

#### Genetic association with gut microbiome

We assessed the genetic overlap between RG GWAS results and those for gut microbiome. GWAS of microbiome profiles were publicly available and downloaded from the https://mibiogen.gcc.rug.nl/<u>[mibiogen.gcc.rug.nl]</u>. For each of the 211 taxa, the corresponding *P*-values for the 143 RG GWAS SNPs and their proxies were extracted.

#### Genetic association with GLP-1 and GIP

We assessed the genetic overlap between RG GWAS results and those for glucagon-like peptide-1 (GLP-1) and gastric inhibitory polypeptide (GIP) measured at 0 and 120 minutes. We extracted the results for the 143 RG signals from the GWAS summary statistics for GLP-1 and GIP^87^.

#### eQTL co-localization analysis

We further performed co-localization analysis using whole blood gene expression-QTL (eQTL) data provided by eQTLGen^39^ and AS20+AST20 meta-analysis results. Only cis-eQTL data from eQTLGen was incorporated to reduce the computational burden. The COLOC2 Bayesian-based method^88^ was used to interrogate the potential co-localization between RG GWAS signals and the genetic control of gene expression. We first extracted the RG GWAS test statistics of all the SNPs within +/-1Mb region around the 143 RG signals. Then, for each RG signal, we matched the eQTLGen results with the RG results and performed COLOC2 analysis evaluating the posterior probability (PP) of five hypotheses for each region: H_0_, no association; H_1_, GWAS association only; H_2_, eQTL association only; H_3_, both GWAS and eQTL association, but not co-localised; and H_4_, both GWAS and eQTL association and co-localised. Only GWAS signals with at least one nearby gene/probe reaching PP (H_4_) ≥ 0.5 were reported.

#### Genetic association with human blood plasma N-glycosylation

We assessed the genetic association between 143 RG signals and 113 human blood plasma N-glycome traits using previously published genome-wide summary association statistics^89^. The description of the analysed traits and details of the association analysis can be found elsewhere^40^. We considered associations to be significant when *P*-value<0.05/113/143=3.09e-6 (after Bonferroni correction). Association was considered as suggestive when *P*-value<10^−4^.

#### Genetic correlation analysis

We investigate the shared genetic component between RG and other traits, including glycaemic ones, by performing genetic correlation analysis using the bivariate LD score regression method (LDSC v1.0.0)^90^. To reduce multiple testing burden, only the GWAS results of the UKBB model AS20 were used. We used GWAS summary statistics available in LDhub^91^ and the Meta-Analysis of Glucose and Insulin-related Traits Consortium (MAGIC) website (https://www.magicinvestigators.org) for several traits including FG/FI^54^, HOMA-B/HOMA-IR^92^. In total, 228 different traits were included in the genetic correlation analysis with RG. We considered *P*≤0.05 as the nominal significant level.

#### MR analysis

We applied a bidirectional two-sample MR strategy to investigate causality between RG and lung function, as well as T2D and lung function using independent genetic variants as instruments. MR can provide estimates of the effect of modifiable exposures on an outcome (e.g. disease) unaffected by classical confounding or reverse causation, whenever randomised clinical trials are not feasible. We looked for evidence for the presence of a causal effect of RG and T2D on two lung function phenotypes; FVC and FEV1 in a two-sample MR setting. Genome-wide summary statistics for the lung function phenotypes were available^93^, involving cohorts from the SpiroMeta consortium and the UKBB study. T2D susceptibility variants and their effects were obtained from the largest-to-date T2D GWAS^4^.

To avoid confounding due to sample overlap, lung function summary statistics used as outcome data were those estimated in the SpiroMeta consortium alone. Similarly, when testing the effect of lung function on RG, RG genetic effects used as outcome data were estimated in all cohorts except UK Biobank. There was no sample overlap between the lung function- and the T2D GWAS, thus allowing the use of T2D effects estimated in all contributing European studies. Genome-wide T2D summary statistics were available from a previous study^3^ to test for the causal effect of lung function on T2D. All analyses were conducted using the R software package TwoSampleMR v0.5.4^94^.

Instrument selection: Independent (established by conditional analyses for both RG and the lung function phenotypes) genome-wide significant (*P*<5×10^−8^) variants were selected as genetic instruments. In total, 143 independent variants were defined for RG by the current study, 424 T2D signals were reported for Europeans by Vujkovic *et al.* and 130/162 independent signals were reported by Shrine *et al.* for FVC and FEV1, respectively. We looked for proxy variants with a minimum r^2^ of 0.8 where the instrumental variant was not present in the outcome data. Palindromic variants with minor allele frequency larger than 45% were excluded to avoid uncertainty when harmonizing effects to the exposure-increasing allele. After filtering, 136 variants were used to instrument RG and 413 variants were available as T2D instruments. For FVC, 125 and 115 variants could be used as instruments in the RG and T2D MR analyses, respectively. For FEV1, 157 and 140 variants served as instruments in the RG and T2D MR analyses, respectively.

Causal effects were estimated using the inverse-variance weighted method, which combines the causal estimates of individual instrumental variants (Wald ratios) in a random-effects meta-analysis^95^. As a sensitivity analysis, we employed MR-Egger regression to obtain causal estimates that are more robust to the inclusion of invalid instruments^96^.

#### PRS analysis

We tested the ability of the RG genetic effects to predict RG, T2D and HbA1c. We compared that to the predictive power of T2D and FG genetic instruments by computing PRS for RG, T2D and FG and assessing their performance in predicting RG, T2D and HbA1c. PRS analyses require base- and target data from independent populations. The base datasets in our analyses were UKBB-only estimates from the present RG GWAS, meta-analysis estimates of 32 studies for T2D^97^ and meta-analysis estimates from the MAGIC for FG^54^. We used the second largest cohort, the Vanderbilt University Medical Centre (VUMC), as our target dataset. PRS construction and model evaluation were done using the software PRSice (v2.2.3)^98^. The PRS for an individual is the summation of the effect (trait-increasing) alleles weighted by the effect size of the SNP taken from the base data. The SNPs in the base data are clumped so that they are largely independent of each other and thus their effects can be summed. To assess predictive power, PRS for RG, T2D and FG were regressed onto the phenotypes of interest (i.e. RG, T2D and HbA1c) providing the coefficient of determination (R^2^) as an estimate for the correlation between the phenotype and the PRS in the VUMC cohort. All models were adjusted for age, four principal components, sex and the cohort-specific batch effect. Since the optimal *P*-value threshold for including SNPs in the PRS is unknown a priori, PRS are calculated over a range of thresholds and regressed onto the phenotype of interest, optimising prediction accordingly. The R^2^ estimates for each trait were derived by subtracting the R^2^ from the null model (*Phenotype ∼ sex + age + 4 principal components + batch)* from the R^2^ from the full model (*Phenotype ∼ PRS + sex + age + 4 principal components + batch*) which contains the PRS at the best predicting P-value threshold.

#### Clustering of the RG signals with results for 45 other phenotypes

We looked up the Z-scores (regression coefficient beta divided by the standard error) of the distinct 143 RG signals in publicly available summary statistics of 45 relevant phenotypes. All variant effects were aligned to the RG risk allele. HapMap2 based summary statistics were imputed using SS-Imp v0.5.5^65^ to minimise missingness. Missing summary statistics values were imputed via mean imputation. The resulting variant-trait association matrix was scaled by the square root of the study’s mean sample size. We used agglomerative hierarchical clustering with Ward’s method to partition the variants into groups by their effects on the considered outcomes. The clustering analysis was performed in R using function hclust() from in-built stats package.

## Data Availability

GWAS summary statistics for RG analyses presented in this manuscript will be deposited on the MAGIC website and will be also be available through the NHGRI-EBI GWAS Catalog.

https://www.magicinvestigators.org/downloads/

https://www.ebi.ac.uk/gwas/downloads/summary-statistics

## Data availability

GWAS summary statistics for RG analyses presented in this manuscript will be deposited on https://www.magicinvestigators.org/downloads/ and will be also be available through the NHGRI-EBI GWAS Catalog https://www.ebi.ac.uk/gwas/downloads/summary-statistics.

## Acknowledgements

### Airwave

The Airwave Health Monitoring Study was funded by the UK Home Office (780-TETRA, 2003-2018) and is currently funded by the MRC and ESRC (MR/R023484/1) with additional funding from the NIHR Imperial College Biomedical Research Centre (BRC) in collaboration with Imperial College NHS Healthcare Trust. We thank all Airwave participants for their contribution to the study.

Personal support: Paul Elliott acknowledges support from the MRC and PHE (MR/L01341X/1, 812 2014-2019) and currently from the MRC for the MRC Centre for Environment and Health 813 (MR/S019669/1). Paul Elliott and Ioanna Tzoulaki are supported by the UK Dementia Research Institute which 814 receives funding from UK DRI Ltd funded by the UK Medical Research Council, Alzheimer’s 815 Society and Alzheimer’s Research UK. Paul Elliott is associate director of the Health Data 816 Research UK London funded by a consortium led by the UK Medical Research Council.

### BRIGHT

This work was funded by the Medical Research Council of Great Britain (grant number: G9521010D). The BRIGHT study is extremely grateful to all the patients who participated in the study and the BRIGHT nursing team. This work formed part of the research themes contributing to the translational research portfolio for the NIHR Barts Cardiovascular Biomedical Research Centre. The funders had no role in study design, data collection and analysis.

### deCODE

We thank participants in deCODE genetic studies whose contribution made this work possible.

### EMIL

EMIL-Cohort, a population-based cohort; the study was approved by the ethical committee of the Chamber of Physicians Baden-Württemberg in the year 2002 (Registration Number 133-02; dates 05.09.2002 and 24.09.2002). We thank Silke Rosinger, Simone Claudi-Boehm, Rosina Sing, Sabine Schilling and Angelika Kurkhaus for technical support.

### EPIC-Norfolk

The EPIC-Norfolk study (https://doi.org/10.22025/2019.10.105.00004) has received funding from the Medical Research Council (MR/N003284/1 and MC-UU_12015/1) and Cancer Research UK (C864/A14136). The genetics work in the EPIC-Norfolk study was funded by the Medical Research Council (MC_PC_13048). We are grateful to all the participants who have been part of the project and to the many members of the study teams at the University of Cambridge who have enabled this research.

### FINRISK87

Support for FUSION was provided by NIH grants R01-DK062370 (to M.B.), R01-DK072193 (to K.L.M.), and intramural project number 1Z01-HG000024 (to F.S.C.). Genome-wide genotyping was conducted by the Johns Hopkins University Genetic Resources Core Facility SNP Center at the Center for Inherited Disease Research (CIDR), with support from CIDR NIH contract no. N01-HG-65403.

### Framingham Heart Study

Also supported by National Institute for Diabetes and Digestive and Kidney Diseases (NIDDK) U01/UM1 DK078616 to Dr. Meigs

### HUNT2/Tromson

The Nord-Trøndelag Health Study (the HUNT study) is a collaboration between HUNT Research Centre (Faculty of Medicine, Norwegian University of Science and Technology NTNU), Nord-Trøndelag County Council, Central Norway Health Authority, and the Norwegian Institute of Public Health. University of Tromsø, Norwegian Research Council (project number 185764).

### InterAct

We thank all EPIC participants and staff and the InterAct Consortium members for their contributions to the study. The InterAct project received funding from the European Union (Integrated Project LSHM-CT-2006-037197 in the Framework Programme 6 of the European Community). We thank staff from the technical, field epidemiology and data teams of the Medical Research Council Epidemiology Unit in Cambridge, UK, for carrying out sample preparation, DNA provision and quality control, genotyping and data handling work.

### KORA F3

The KORA research platform (KORA, Cooperative Research in the Region of Augsburg) was initiated and financed by the Helmholtz Zentrum München–German Research Center for Environment and Health, which is funded by the German Federal Ministry of Education and Research and by the state of Bavaria. Furthermore, part of this work was supported by the German National Genome Research Network (NGFN) and the Munich Center of Health Sciences (MC Health) as part of LMUinnovativ.

### Brisbane Adolescent Twin Study / SSAGA-NAG adult cohort

Genotyping and phenotyping were supported by the Australian National Health and Medical Research Council (389891, 389892, 496739), the EU 5th Framework Programme GenomEUtwin Project (QLG2-CT-2002-01254) and the U.S. National Institutes of Health (AA07535, AA13320, AA13321, AA13326, AA14041, DA12854). B.B. and G.W.M. are supported by National Health and Medical Research Council (NHMRC) Fellowship Schemes. Participants gave informed consent and the studies were approved by appropriate institutional review boards.

### PPP-Botnia/Malmö Diet Cancer

Personal support: Emma Ahlqvist was funded by grants from the Swedish Research Council (2017-02688, 2020-02191).

### PROCARDIS

PROCARDIS was supported by the European Community Sixth Framework Program (LSHM-CT-2007-037273), AstraZeneca, the British Heart Foundation, the Wellcome Trust (Contract No. 075491/Z/04), the Swedish Research Council, the Knut and Alice Wallenberg Foundation, the Swedish Heart-Lung Foundation, the Torsten and Ragnar Söderberg Foundation, the Strategic Cardiovascular and Diabetes Programs of Karolinska Institutet and Stockholm County Council, the Foundation for Strategic Research and the Stockholm County Council. Ethical permission was granted by the local ethical board for each centre.

Personal support: Anuj Goel has received support from the BHF, European Commission [LSHM-CT-2007-037273, HEALTH-F2-2013-601456] and TriPartite Immunometabolism Consortium [TrIC]-NovoNordisk Foundation [NNF15CC0018486]. Hugh Watkins has received support from Wellcome Trust core awards (090532/Z/09/Z, 203141/Z/16/Z) and is member of the Oxford BHF Centre for Research Excellence (RE/13/1/30181). Rona J Strawbridge is supported by a UKRI Innovation-HDR-UK Fellowship (MR/S003061/1).

### Rotterdam Study

The generation and management of GWAS genotype data for the Rotterdam Study is supported by the Netherlands Organisation of Scientific Research NWO Investments (nr. 175.010.2005.011, 911-03-012). This study is funded by the Research Institute for Diseases in the Elderly (014-93-015; RIDE2), the Netherlands Genomics Initiative (NGI)/Netherlands Organisation for Scientific Research (NWO) project nr. 050-060-810. We thank Pascal Arp, Mila Jhamai, Marijn Verkerk, Lizbeth Herrera and Marjolein Peters for their help in creating the GWAS database, and Karol Estrada and Maksim V. Struchalin for their support in creation and analysis of imputed data. The Rotterdam Study is funded by Erasmus Medical Center and Erasmus University, Rotterdam, Netherlands Organization for the Health Research and Development (ZonMw), the Research Institute for Diseases in the Elderly (RIDE), the Ministry of Education, Culture and Science, the Ministry for Health, Welfare and Sports, the European Commission (DG XII), and the Municipality of Rotterdam. The authors are grateful to the study participants, the staff from the Rotterdam Study and the participating general practitioners and pharmacists.

**The Section of Endocrinology and Investigative Medicine** is funded by grants from the MRC, BBSRC, NIHR, and is supported by the NIHR Biomedical Research Centre Funding Scheme. The views expressed are those of the author(s) and not necessarily those of the any of the funders, the NHS, the NIHR or the Department of Health.

### UK Biobank

We thank UK Biobank for data availability, project number 37685.

Personal support: Marika A Kaakinen, Anna Ulrich, Zhanna Balkhiyarova and Inga Prokopenko are in part funded by the European Union’s Horizon 2020 research and innovation programme LONGITOOLS, H2020-SC1-2019-874739. Marika A Kaakinen is also funded by the European Foundation for the Study of Diabetes (EFSD) Albert Renold Travel Fellowship.

### Vanderbilt

The samples and/or dataset(s) used for the analyses described were obtained from Vanderbilt University Medical Center’s BioVU which is supported by numerous sources: institutional funding, private agencies, and federal grants. These include the NIH funded Shared Instrumentation Grant S10OD017985 and S10RR025141; and CTSA grants UL1TR002243, UL1TR000445, and UL1RR024975. Genomic data are also supported by investigator-led projects that include U01HG004798, R01NS032830, RC2GM092618, P50GM115305, U01HG006378, U19HL065962, R01HD074711; and additional funding sources listed at https://victr.vumc.org/biovu-funding/”.

Personal support: NIH Grants R01 HL146588 and R01 HL146588-01S1.

### Individual acknowledgements

**Alejandra Tomas** was supported by MRC project grant MR/R010676/1 and grants from Diabetes UK and the European Federation for the Study of Diabetes.

**Alessia David** is funded by Wellcome Trust grant 104955/Z/14/Z.

**Ben Jones** is supported by an Imperial Post-CCT Post-Doctoral Fellowship, MRC project grant MR/R010676/1, and grants from the European Federation for the Study of Diabetes, Academy of Medical Sciences, Imperial NIHR Biomedical Research Centre, Engineering and Physical Sciences Research Council, Society for Endocrinology and British Society for Neuroendocrinology.

**Christopher Reynolds** is a Royal Society Industry Fellow.

**Inês Barroso** was funded by an “Expanding excellence in England” award from Research England.

**Inga Prokopenko** is funded by the World Cancer Research Fund (WCRF UK) and World Cancer Research Fund International (2017/1641), the Wellcome Trust (WT205915), the European Union’s Horizon 2020 research and innovation programme (DYNAhealth, project number 633595) the LONGITOOLS, H2020-SC1-2019-874739 and the Royal Society (IEC\R2\181075).

**Mark McCarthy** was a Wellcome Investigator and an NIHR Senior Investigator (Niddk. U01-DK105535, Wellcome: 090532, 098381, 106130, 203141, 212259).

**Patrick M Sexton and Denise Wootten**: The work was supported by NHMRC Ideas #1184726 and NHMRC program grant #1150083. Patrick M Sexton is a NHMRC Senior Principal Research Fellow (#1154434) and Denise Wootten is an NHMRC Senior Research Fellow (#1155302).

**Vasiliki Lagou** was supported by a fellowship from the Research Foundation-Flanders (FWO).

**Victoria Salem** is the recipient of a Diabetes UK Harry Keen Clinician Scientist Fellowship.

**Sharapov Sodbo** was supported by a grant from the Russian Science Foundation (RSF) No. 19-15-00115.

**Tricia M Tan** acknowledges support from the National Institute for Health Research (NIHR) Imperial Biomedical Research Centre (BRC) Funding Scheme.

## Author contributions

**First author**: V.L., L.J., A.U., **Central analysis and writing group**: V.L., L.J., A.U., L.Z., K.G., Z.B., A.F., L.M., **Additional analyses junior lead**: S.C., P.T., S.S., A.David, R.M., R.R., E.Ahlqvist, T.M.T., A.T., V.Salem, **GWAS cohort analyst**: G.T., Η.G., E.E., B.B., R.S., A.I., J.Z., S.M.W., T.J., C.G., H.G., C.M., M.M., R.J.S., A.G., D.R., J.D., Y.S.A., M.A.K., **Metabochip cohort analyst**: E.Albrecht, A.U.J., H.M.S., **Cohort sample collection, genotyping, phenotyping or additional analysis**: I.R.C., F.E., V.Steinthorsdottir, A.G.U., P.B.M., M.J.B., S.J., O.H., B.T., K.H., T.W., K.L.M., **Metabochip cohort PI**: W.Kratzer, H.M., W.Koenig, B.O.B., **GWAS cohort PI**: J.T., M.B., J.C.F., A.Hamsten, H.Watkins, I.N., H.Wichmann, M.J.C., K.K., C.v.D., A.Hofman, N.J.W., C.L., J.B.W., N.G.M., G.M., I.T., P.E., U.T., K.S., E.L.B., J.B.M., **Additional analyses senior**: P.M.S., D.W., L.G., G.D., A.Demirkan, T.H.P., C.A.R., **Other senior author (analysis and writing group)**: Y.S.A., **Other senior author (writing)**: I.B., C.S., M.I.M., P.F., J.D., J.B.M., **Senior author**: M.A.K., B.J., I.P.

## Competing interests

**Alejandra Tomas** has received grant funding from Sun Pharmaceuticals.

**Ivan R Corrêa, Jr** is an employee of New England Biolabs, Inc., a manufacturer and vendor of reagents for life science research.

**Mark J Caulfield** is Chief Scientist for Genomics England, a UK Government company.

The views expressed in this article are those of the author(s) and not necessarily those of the NHS, the NIHR, or the Department of Health. **Mark McCarthy** has served on advisory panels for Pfizer, NovoNordisk and Zoe Global, has received honoraria from Merck, Pfizer, Novo Nordisk and Eli Lilly, and research funding from Abbvie, Astra Zeneca, Boehringer Ingelheim, Eli Lilly, Janssen, Merck, NovoNordisk, Pfizer, Roche, Sanofi Aventis, Servier, and Takeda. As of June 2019, MMcC is an employee of Genentech, and a holder of Roche stock.

**Patrick M Sexton** receives grant funding from Laboratoires Servier.

**Toby Johnson** is now a GSK employee.

**Yurii S Aulchenko** is owner of Maatschap PolyOmica and PolyKnomics BV, private organizations providing services, research and development in the field of computational and statistical, quantitative and computational (gen)omics.

## Notes

### Author Declarations

North West - Haydock Research Ethics Committee, UK

## References

1. Santos, R.L. et al. Heritability of fasting glucose levels in a young genetically isolated population. Diabetologia 49, 667–72 (2006).

2. Almgren, P. et al. Heritability and familiality of type 2 diabetes and related quantitative traits in the Botnia Study. Diabetologia 54, 2811–9 (2011).

3. Scott, R.A. et al. An Expanded Genome-Wide Association Study of Type 2 Diabetes in Europeans. Diabetes 66, 2888–2902 (2017).

4. Vujkovic, M. et al. Discovery of 318 new risk loci for type 2 diabetes and related vascular outcomes among 1.4 million participants in a multi-ancestry meta-analysis. Nat Genet 52, 680–691 (2020).

5. Chen, J. et al. The Trans-Ancestral Genomic Architecture of Glycaemic Traits. bioRxiv, 2020.07.23.217646 (2020).

6. Dimas, A.S. et al. Impact of type 2 diabetes susceptibility variants on quantitative glycemic traits reveals mechanistic heterogeneity. Diabetes 63, 2158–71 (2014).

7. Ingelsson, E. et al. Detailed physiologic characterization reveals diverse mechanisms for novel genetic Loci regulating glucose and insulin metabolism in humans. Diabetes 59, 1266–75 (2010).

8. Scott, R.A. et al. Large-scale association analyses identify new loci influencing glycemic traits and provide insight into the underlying biological pathways. Nat Genet 44, 991–1005 (2012).

9. Deng, Y.N., Xia, Z., Zhang, P., Ejaz, S. & Liang, S. Transcription Factor RREB1: from Target Genes towards Biological Functions. Int J Biol Sci 16, 1463–1473 (2020).

10. Piccand, J. et al. Rfx6 maintains the functional identity of adult pancreatic beta cells. Cell Rep 9, 2219–32 (2014).

11. Tsuboi, T. et al. Glucagon-like peptide-1 mobilizes intracellular Ca2+ and stimulates mitochondrial ATP synthesis in pancreatic MIN6 beta-cells. Biochem J 369, 287–99 (2003).

12. Bosma, K.J. et al. Pancreatic islet beta cell-specific deletion of G6pc2 reduces fasting blood glucose. J Mol Endocrinol 64, 235–248 (2020).

13. Rutter, G.A., Georgiadou, E., Martinez-Sanchez, A. & Pullen, T.J. Metabolic and functional specialisations of the pancreatic beta cell: gene disallowance, mitochondrial metabolism and intercellular connectivity. Diabetologia 63, 1990–1998 (2020).

14. Hara, K. et al. Genome-wide association study identifies three novel loci for type 2 diabetes. Hum Mol Genet 23, 239–46 (2014).

15. Morris, A.P. et al. Large-scale association analysis provides insights into the genetic architecture and pathophysiology of type 2 diabetes. Nat Genet 44, 981–90 (2012).

16. Mahajan, A. et al. Identification and functional characterization of G6PC2 coding variants influencing glycemic traits define an effector transcript at the G6PC2-ABCB11 locus. PLoS Genet 11, e1004876 (2015).

17. Pullen, T.J. & Rutter, G.A. Roles of lncRNAs in pancreatic beta cell identity and diabetes susceptibility. Front Genet 5, 193 (2014).

18. Benonisdottir, S. et al. Sequence variants associating with urinary biomarkers. Hum Mol Genet 28, 1199–1211 (2019).

19. Teumer, A. et al. Genome-wide association meta-analyses and fine-mapping elucidate pathways influencing albuminuria. Nat Commun 10, 4130 (2019).

20. Wuttke, M. et al. A catalog of genetic loci associated with kidney function from analyses of a million individuals. Nat Genet 51, 957–972 (2019).

21. Spracklen, C.N. et al. Identification of type 2 diabetes loci in 433,540 East Asian individuals. Nature 582, 240–245 (2020).

22. Wessel, J. et al. Low-frequency and rare exome chip variants associate with fasting glucose and type 2 diabetes susceptibility. Nat Commun 6, 5897 (2015).

23. Tomkin, G.H. Treatment of type 2 diabetes, lifestyle, GLP1 agonists and DPP4 inhibitors. World J Diabetes 5, 636–50 (2014).

24. Koole, C. et al. Polymorphism and ligand dependent changes in human glucagon-like peptide-1 receptor (GLP-1R) function: allosteric rescue of loss of function mutation. Mol Pharmacol 80, 486–97 (2011).

25. Wan, Q. et al. Mini G protein probes for active G protein-coupled receptors (GPCRs) in live cells. J Biol Chem 293, 7466–7473 (2018).

26. Jones, B. et al. Targeting GLP-1 receptor trafficking to improve agonist efficacy. Nat Commun 9, 1602 (2018).

27. Deganutti, G. et al. Dynamics of GLP-1R peptide agonist engagement are correlated with kinetics of G protein activation. bioRxiv, 2021.03.10.434902 (2021).

28. Venkatakrishnan, A.J. et al. Diverse GPCRs exhibit conserved water networks for stabilization and activation. Proc Natl Acad Sci U S A 116, 3288–3293 (2019).

29. Yuan, S., Filipek, S., Palczewski, K. & Vogel, H. Activation of G-protein-coupled receptors correlates with the formation of a continuous internal water pathway. Nat Commun 5, 4733 (2014).

30. Zhao, P. et al. Activation of the GLP-1 receptor by a non-peptidic agonist. Nature 577, 432–436 (2020).

31. Karczewski, K.J. et al. The mutational constraint spectrum quantified from variation in 141,456 humans. Nature 581, 434–443 (2020).

32. Hauser, A.S. et al. Pharmacogenomics of GPCR Drug Targets. Cell 172, 41–54 e19 (2018).

33. Sorli, C. et al. Efficacy and safety of once-weekly semaglutide monotherapy versus placebo in patients with type 2 diabetes (SUSTAIN 1): a double-blind, randomised, placebo-controlled, parallel-group, multinational, multicentre phase 3a trial. Lancet Diabetes Endocrinol 5, 251–260 (2017).

34. Pers, T.H. et al. Biological interpretation of genome-wide association studies using predicted gene functions. Nat Commun 6, 5890 (2015).

35. Timshel, P.N., Thompson, J.J. & Pers, T.H. Genetic mapping of etiologic brain cell types for obesity. Elife 9(2020).

36. Ding, Q. et al. Genome-wide meta-analysis associates GPSM1 with type 2 diabetes, a plausible gene involved in skeletal muscle function. J Hum Genet 65, 411–420 (2020).

37. Kurilshikov, A. et al. Genetics of human gut microbiome composition. bioRxiv, 2020.06.26.173724 (2020).

38. Lopera-Maya, E.A. et al. Effect of host genetics on the gut microbiome in 7,738 participants of the Dutch Microbiome Project. bioRxiv, 2020.12.09.417642 (2020).

39. Võsa, U. et al. Unraveling the polygenic architecture of complex traits using blood eQTL metaanalysis. bioRxiv, 447367 (2018).

40. Sharapov, S.Z. et al. Defining the genetic control of human blood plasma N-glycome using genome-wide association study. Hum Mol Genet 28, 2062–2077 (2019).

41. Clerc, F. et al. Human plasma protein N-glycosylation. Glycoconj J 33, 309–43 (2016).

42. Novokmet, M. et al. Changes in IgG and total plasma protein glycomes in acute systemic inflammation. Sci Rep 4, 4347 (2014).

43. Schmidt, M.I. et al. Markers of inflammation and prediction of diabetes mellitus in adults (Atherosclerosis Risk in Communities study): a cohort study. Lancet 353, 1649–52 (1999).

44. Dotz, V. et al. Plasma protein N-glycan signatures of type 2 diabetes. Biochim Biophys Acta Gen Subj 1862, 2613–2622 (2018).

45. Keser, T. et al. Increased plasma N-glycome complexity is associated with higher risk of type 2 diabetes. Diabetologia 60, 2352–2360 (2017).

46. Wittenbecher, C. et al. Plasma N-Glycans as Emerging Biomarkers of Cardiometabolic Risk: A Prospective Investigation in the EPIC-Potsdam Cohort Study. Diabetes Care 43, 661–668 (2020).

47. Johswich, A. et al. N-glycan remodeling on glucagon receptor is an effector of nutrient sensing by the hexosamine biosynthesis pathway. J Biol Chem 289, 15927–41 (2014).

48. Lemmers, R.F.H. et al. IgG glycan patterns are associated with type 2 diabetes in independent European populations. Biochim Biophys Acta Gen Subj 1861, 2240–2249 (2017).

49. Liu, D. et al. Ischemic stroke is associated with the pro-inflammatory potential of N-glycosylated immunoglobulin G. J Neuroinflammation 15, 123 (2018).

50. Kopf, S. et al. Breathlessness and Restrictive Lung Disease: An Important Diabetes-Related Feature in Patients with Type 2 Diabetes. Respiration 96, 29–40 (2018).

51. Sonoda, N. et al. A prospective study of the impact of diabetes mellitus on restrictive and obstructive lung function impairment: The Saku study. Metabolism 82, 58–64 (2018).

52. Abdi, A., Jalilian, M., Sarbarzeh, P.A. & Vlaisavljevic, Z. Diabetes and COVID-19: A systematic review on the current evidences. Diabetes Res Clin Pract 166, 108347 (2020).

53. Zhu, L. et al. Association of Blood Glucose Control and Outcomes in Patients with COVID-19 and Pre-existing Type 2 Diabetes. Cell Metab 31, 1068–1077 e3 (2020).

54. Lagou, V. et al. Sex-dimorphic genetic effects and novel loci for fasting glucose and insulin variability. Nat Commun 12, 24 (2021).

55. Marullo, L., El-Sayed Moustafa, J.S. & Prokopenko, I. Insights into the genetic susceptibility to type 2 diabetes from genome-wide association studies of glycaemic traits. Curr Diab Rep 14, 551 (2014).

56. Mingrone, G. et al. Metabolic surgery versus conventional medical therapy in patients with type 2 diabetes: 10-year follow-up of an open-label, single-centre, randomised controlled trial. Lancet 397, 293–304 (2021).

57. Whang, A., Nagpal, R. & Yadav, H. Bi-directional drug-microbiome interactions of anti-diabetics. EBioMedicine 39, 591–602 (2019).

58. Voight, B.F. et al. The metabochip, a custom genotyping array for genetic studies of metabolic, cardiovascular, and anthropometric traits. PLoS Genet 8, e1002793 (2012).

59. Li, Y., Willer, C., Sanna, S. & Abecasis, G. Genotype imputation. Annu Rev Genomics Hum Genet 10, 387–406 (2009).

60. Marchini, J., Howie, B., Myers, S., McVean, G. & Donnelly, P. A new multipoint method for genome-wide association studies by imputation of genotypes. Nat Genet 39, 906–13 (2007).

61. Fuchsberger, C., Abecasis, G.R. & Hinds, D.A. minimac2: faster genotype imputation. Bioinformatics 31, 782–4 (2015).

62. Loh, P.R., Kichaev, G., Gazal, S., Schoech, A.P. & Price, A.L. Mixed-model association for biobank-scale datasets. Nat Genet 50, 906–908 (2018).

63. Loh, P.R. et al. Efficient Bayesian mixed-model analysis increases association power in large cohorts. Nat Genet 47, 284–90 (2015).

64. Genomes Project, C., et al. A global reference for human genetic variation. Nature 526, 68–74 (2015).

65. Rueger, S., McDaid, A. & Kutalik, Z. Evaluation and application of summary statistic imputation to discover new height-associated loci. PLoS Genet 14, e1007371 (2018).

66. Willer, C.J., Li, Y. & Abecasis, G.R. METAL: fast and efficient meta-analysis of genomewide association scans. Bioinformatics 26, 2190–1 (2010).

67. Magi, R. & Morris, A.P. GWAMA: software for genome-wide association meta-analysis. BMC Bioinformatics 11, 288 (2010).

68. Chang, C.C. et al. Second-generation PLINK: rising to the challenge of larger and richer datasets. Gigascience 4, 7 (2015).

69. Yang, J. et al. Conditional and joint multiple-SNP analysis of GWAS summary statistics identifies additional variants influencing complex traits. Nat Genet 44, 369–75, S1-3 (2012).

70. Fang, Z. et al. The Influence of Peptide Context on Signaling and Trafficking of Glucagon-like Peptide-1 Receptor Biased Agonists. ACS Pharmacol Transl Sci 3, 345–360 (2020).

71. Fang, Z. et al. Ligand-Specific Factors Influencing GLP-1 Receptor Post-Endocytic Trafficking and Degradation in Pancreatic Beta Cells. Int J Mol Sci 21(2020).

72. Ward, R.J., Alvarez-Curto, E. & Milligan, G. Using the Flp-In T-Rex system to regulate GPCR expression. Methods Mol Biol 746, 21–37 (2011).

73. Peng, T. et al. A BaSiC tool for background and shading correction of optical microscopy images. Nat Commun 8, 14836 (2017).

74. Jaccard, N. et al. Automated method for the rapid and precise estimation of adherent cell culture characteristics from phase contrast microscopy images. Biotechnol Bioeng 111, 504–17 (2014).

75. Kenakin, T. A Scale of Agonism and Allosteric Modulation for Assessment of Selectivity, Bias, and Receptor Mutation. Mol Pharmacol 92, 414–424 (2017).

76. Harvey, M.J., Giupponi, G. & Fabritiis, G.D. ACEMD: Accelerating Biomolecular Dynamics in the Microsecond Time Scale. J Chem Theory Comput 5, 1632–9 (2009).

77. Cuzzolin, A., Deganutti, G., Salmaso, V., Sturlese, M. & Moro, S. AquaMMapS: An Alternative Tool to Monitor the Role of Water Molecules During Protein-Ligand Association. ChemMedChem 13, 522–531 (2018).

78. Wakefield, J. A Bayesian measure of the probability of false discovery in genetic epidemiology studies. Am J Hum Genet 81, 208–27 (2007).

79. Pers, T.H., Timshel, P. & Hirschhorn, J.N. SNPsnap: a Web-based tool for identification and annotation of matched SNPs. Bioinformatics 31, 418–20 (2015).

80. Genomes Project, C. et al. A map of human genome variation from population-scale sequencing. Nature 467, 1061–73 (2010).

81. Tabula Muris, C., et al. Single-cell transcriptomics of 20 mouse organs creates a Tabula Muris. Nature 562, 367–372 (2018).

82. Timshel, P.N., Thompson, J.J. & Pers, T.H. Mapping heritability of obesity by brain cell types. bioRxiv, 2020.01.27.920033 (2020).

83. Barbeira, A.N. et al. Exploring the phenotypic consequences of tissue specific gene expression variation inferred from GWAS summary statistics. Nat Commun 9, 1825 (2018).

84. Gamazon, E.R. et al. A gene-based association method for mapping traits using reference transcriptome data. Nat Genet 47, 1091–8 (2015).

85. Delaneau, O., Marchini, J., Genomes Project, C. & Genomes Project, C. Integrating sequence and array data to create an improved 1000 Genomes Project haplotype reference panel. Nat Commun 5, 3934 (2014).

86. Iotchkova, V. et al. Discovery and refinement of genetic loci associated with cardiometabolic risk using dense imputation maps. Nat Genet 48, 1303–1312 (2016).

87. Almgren, P. et al. Genetic determinants of circulating GIP and GLP-1 concentrations. JCI Insight 2(2017).

88. Dobbyn, A. et al. Landscape of Conditional eQTL in Dorsolateral Prefrontal Cortex and Co-localization with Schizophrenia GWAS. Am J Hum Genet 102, 1169–1184 (2018).

89. Sharapov, S., et al. Genome-wide association summary statistics for human blood plasma glycome. (Zenodo, 2018).

90. Finucane, H.K. et al. Partitioning heritability by functional annotation using genome-wide association summary statistics. Nat Genet 47, 1228–35 (2015).

91. Zheng, J. et al. LD Hub: a centralized database and web interface to perform LD score regression that maximizes the potential of summary level GWAS data for SNP heritability and genetic correlation analysis. Bioinformatics 33, 272–279 (2017).

92. Fedko, I.O. et al. Genetics of fasting indices of glucose homeostasis using GWIS unravels tight relationships with inflammatory markers. bioRxiv, 496802 (2018).

93. Shrine, N. et al. New genetic signals for lung function highlight pathways and chronic obstructive pulmonary disease associations across multiple ancestries. Nat Genet 51, 481–493 (2019).

94. Hemani, G. et al. The MR-Base platform supports systematic causal inference across the human phenome. Elife 7(2018).

95. Burgess, S. et al. Guidelines for performing Mendelian randomization investigations. Wellcome Open Res 4, 186 (2019).

96. Bowden, J., Davey Smith, G. & Burgess, S. Mendelian randomization with invalid instruments: effect estimation and bias detection through Egger regression. Int J Epidemiol 44, 512–25 (2015).

97. Mahajan, A. et al. Fine-mapping type 2 diabetes loci to single-variant resolution using high-density imputation and islet-specific epigenome maps. Nat Genet 50, 1505–1513 (2018).

98. Choi, S.W. & O’Reilly, P.F. PRSice-2: Polygenic Risk Score software for biobank-scale data. Gigascience 8(2019).

