## Supplementary Figures for "Random glucose GWAS in 493,036 individuals provides insights into diabetes pathophysiology, complications and treatment stratification"

**Supplementary Figure 1** (**a**) RG trait models tested. The models were labelled according to covariates included and RG cut-offs used. Individuals were included based on two RG cut-offs: <20 mmol/l (20) to account for the effect of extreme RG values and <11.1 mmol/l (11), which is an established threshold for T2D diagnosis. Hence, Model 1 – AS20 refers to adjusted for Age and Sex, using a cut-off of <20 mmol/l and so forth. (**b-e**) Sensitivity plots showing the correlations between association analyses beta coefficients and Z-scores from RG models in UKBB. For panel c, 4,138 individuals were excluded based on HbA1c≥ 6.5%, in addition to the self-reported or diagnosed T2D cases. Variants with a heterogeneity *P*-value ≤ 0.05 (beta-coefficient plot) or a Z-score difference between the two models compared >3 (Z-score plots) are annotated. (**f**) An enrichment plot showing the effect of RG signals (AS20+AST20 model) on T2D. RG and T2D effect sizes are plotted along the Y and X axes, respectively. Point size and colour are proportional to the statistical significance of the variant for T2D, with red indicating higher and blue lower significance, respectively. The dashed line represents the line of best fit. Variants with T2D *P*-value in the lowest decile are labelled. (**g**) An enrichment plot showing the effects of RG signals (AS20+AST20 model) on HOMA-B and HOMA-IR. The effect sizes on HOMA-B and HOMA-IR are plotted along the Y and X axes, respectively. Point size and colour are proportional to the significance of the variant either in HOMA-B or HOMA-IR, depending on which trait has the smaller *P* value. Red colour indicates higher and blue lower statistical significance, respectively. Variants with a *P*-value < $5x10^{-6}$ are labelled.

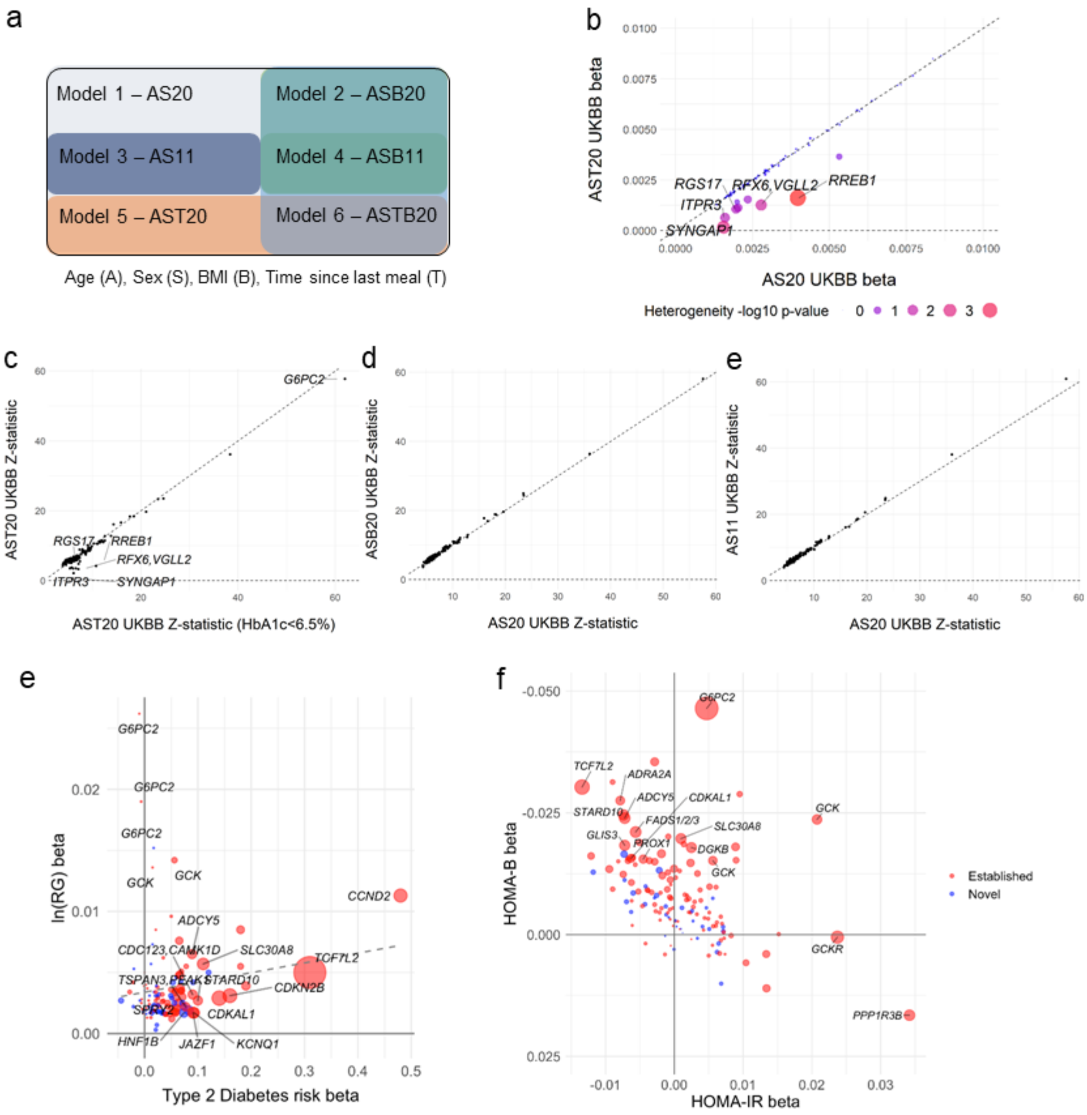

**Supplementary Figure 2** Enrichment plots showing the effect of RG signals (AS20+AST20 model) on glycaemic and respiratory-related phenotypes. (**a**) HbA1c, (**b**) fasting glucose, (**c**) fasting insulin, (**d**) T2D, (**e**) Forced Expiratory Volume in One Second (FEV1), (**f**) Forced Vital Capacity (FVC), (**g**) FEV1/FVC, (**h**) lung cancer and (**i**) squamous cell lung cancer. RG and other phenotype effect sizes are plotted along the Y and X axes, respectively. Point size and colour are proportional to the significance of the variant in each phenotype, with red indicating higher and blue lower significance, respectively. The dashed line represents the line of best fit. Variants with *P*-values in the lowest decile are labelled.

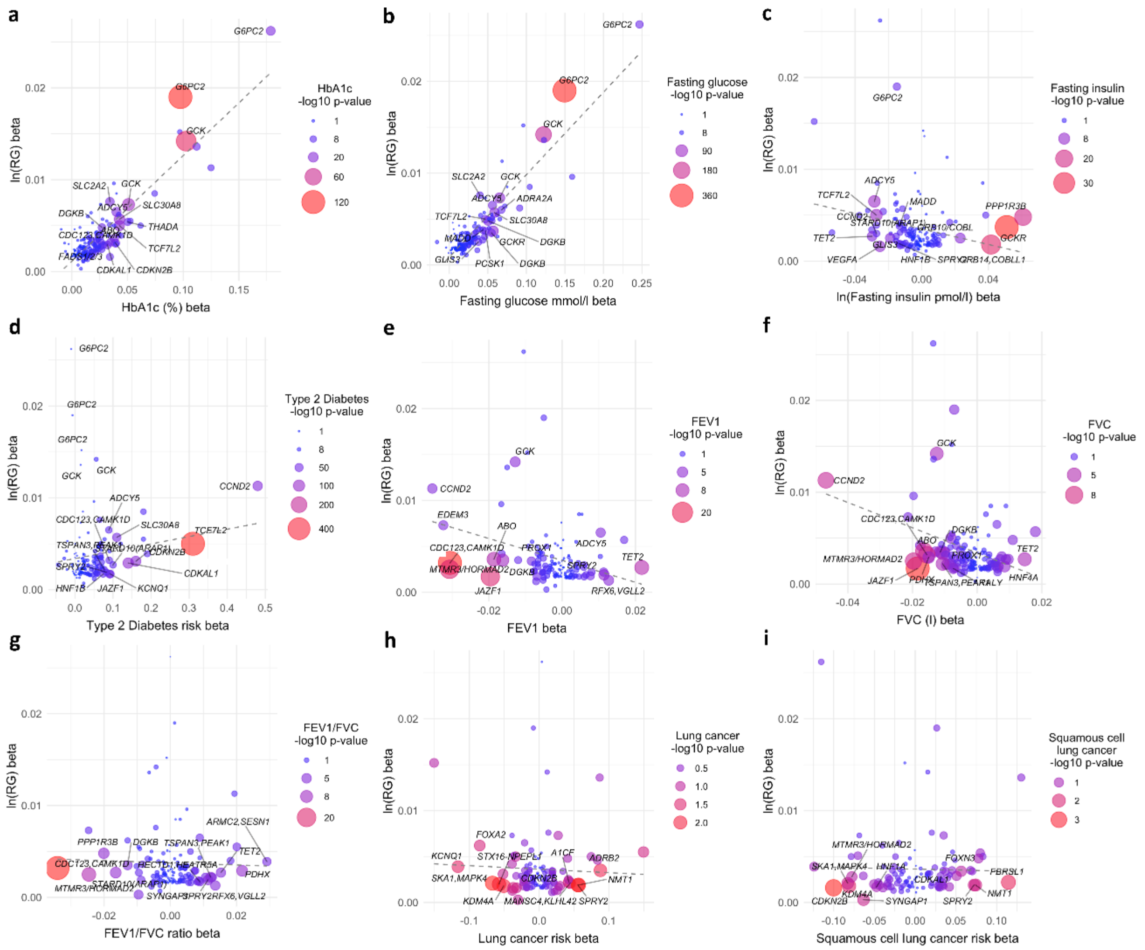

**Supplementary Figure 3** LocusZoom plots of common variants at (**a**) *GCKR*, (**b**) *TET2*, (**c**) *RREB1*, (**d**) *NMT1* (**e**) *WIPI1* loci and low-frequency coding variants at (**f**) *EDEM3*, (**g**) *NEUROD1* and (**h**) *GLP1R* loci.

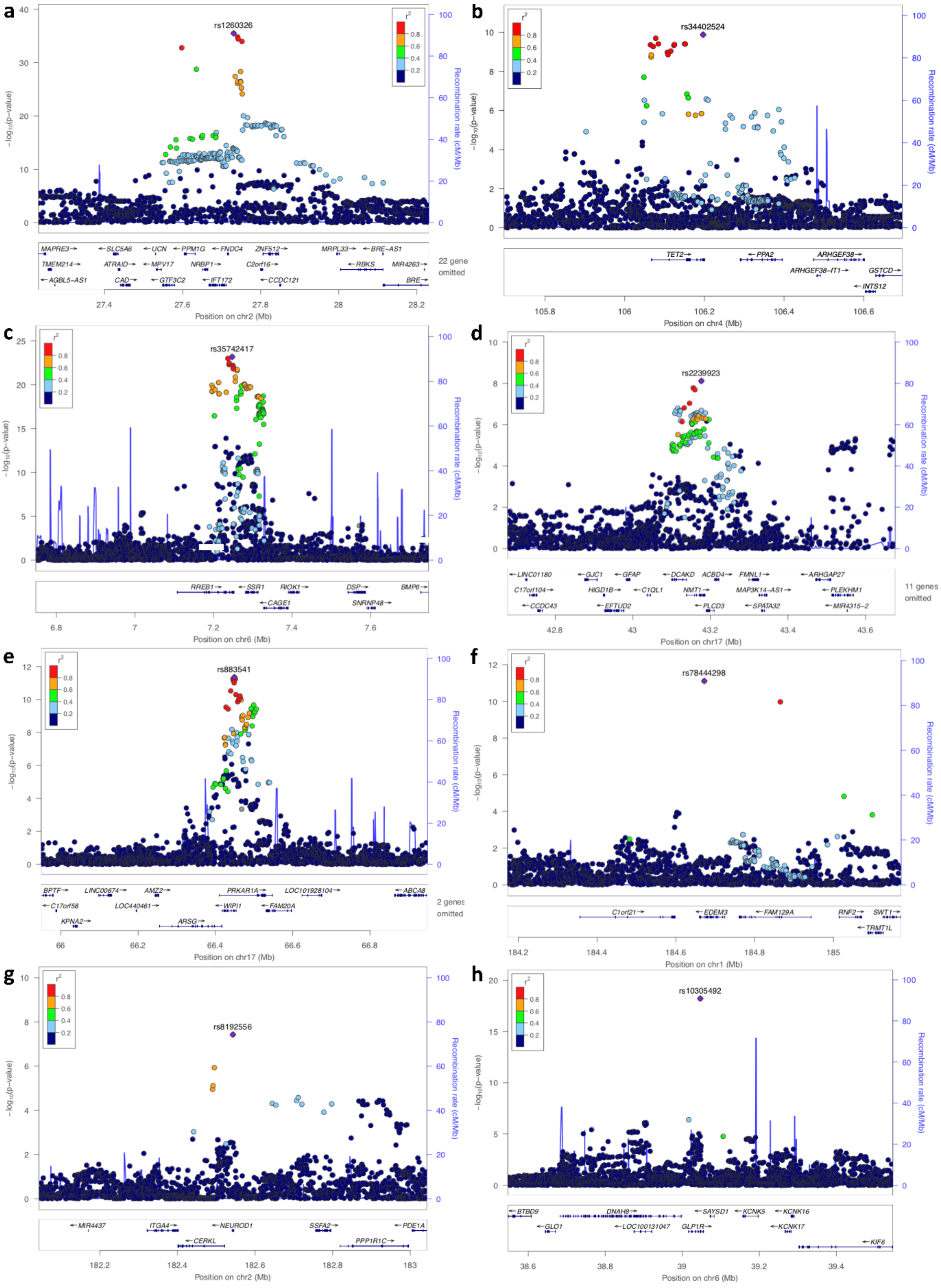

**Supplementary Figure 4** Epigenetic annotation of the RG GWAS results using GARFIELD.

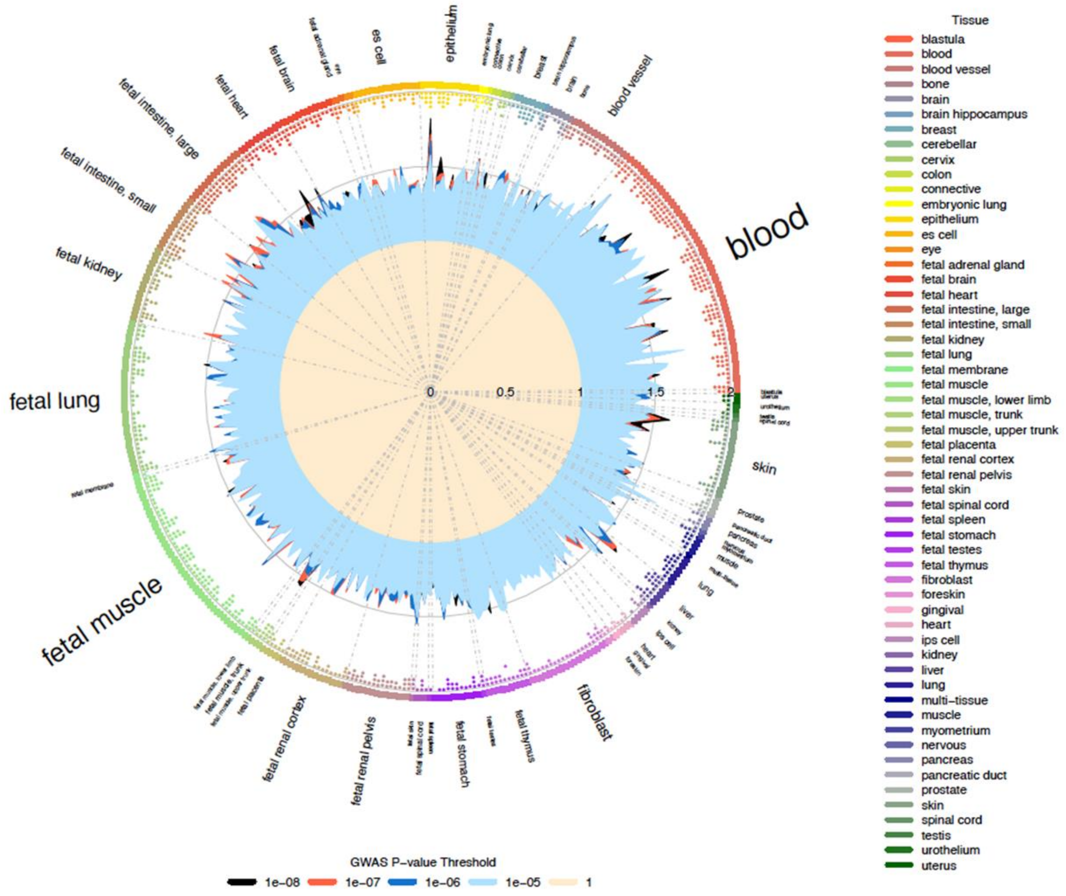

**Supplementary Figure 5** (**a**) Cluster analysis of effects (as Z-scores) of the distinct 143 RG signals on 45 relevant phenotypes (Supplementary Table 23). All variant effects were aligned to the RG risk allele. (**b-d**) Scatter plots of the standardized allelic effect estimates for selected trait pairs. In each scatter plot loci were assigned to thr groups defined from the cluster analysis and highlighted by different colors; (**b**): Corrected Insulin Response (CIR) vs. Type 2 Diabetes (T2D) (clusters 1a/b related to metabolic sydrome); (**c**) Glycated haemoglobin (HbA1c) vs. Inflammatory bowel disease (IBD) (cluster 2a) highlights the effects of loci with a protective role in IBD, and (**d**) Plasminogen activator inhibitor-1 (PAI-1) vs. CIR (cluster 3) highlights loci linked to insulin secretion defects.

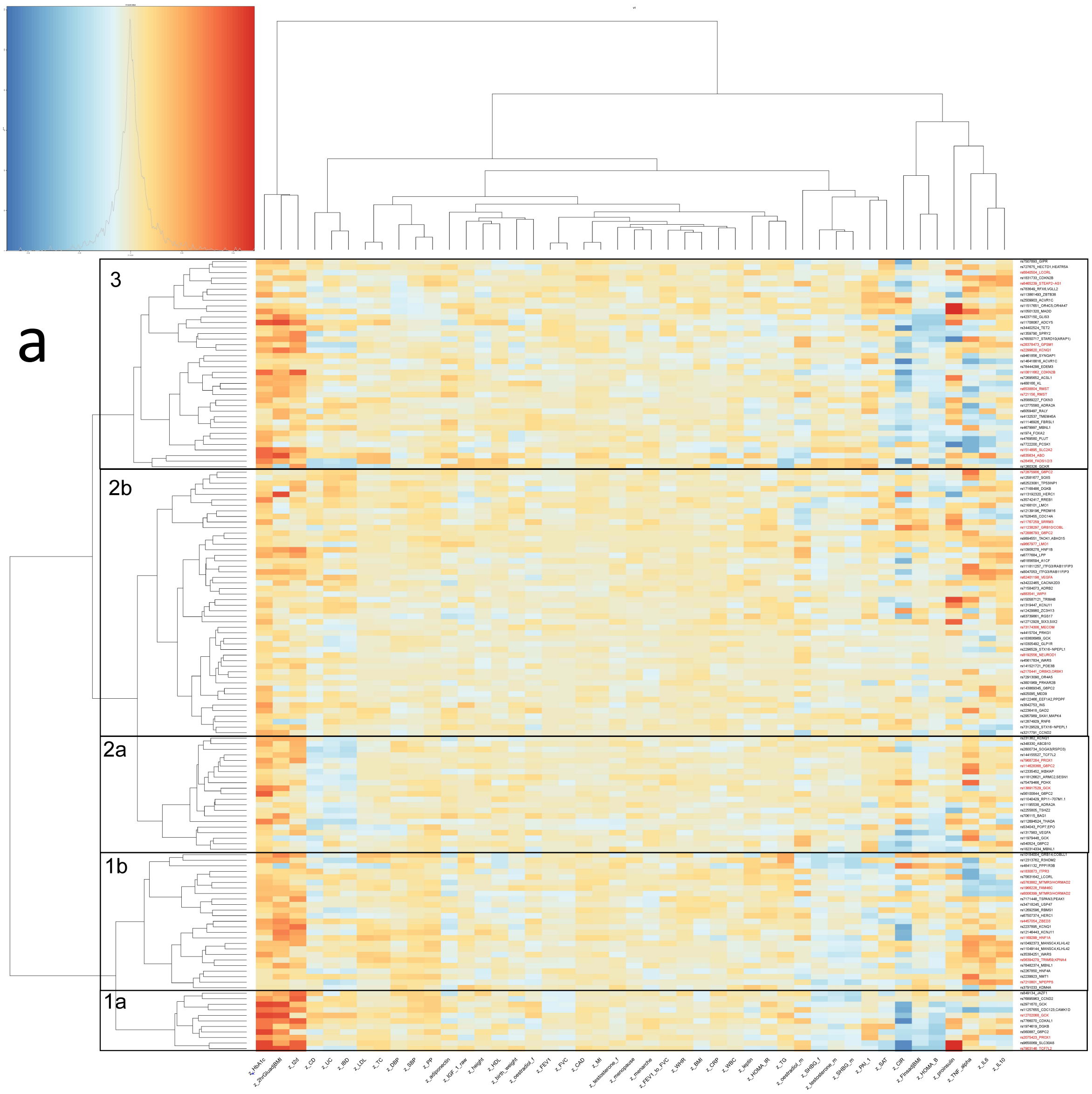

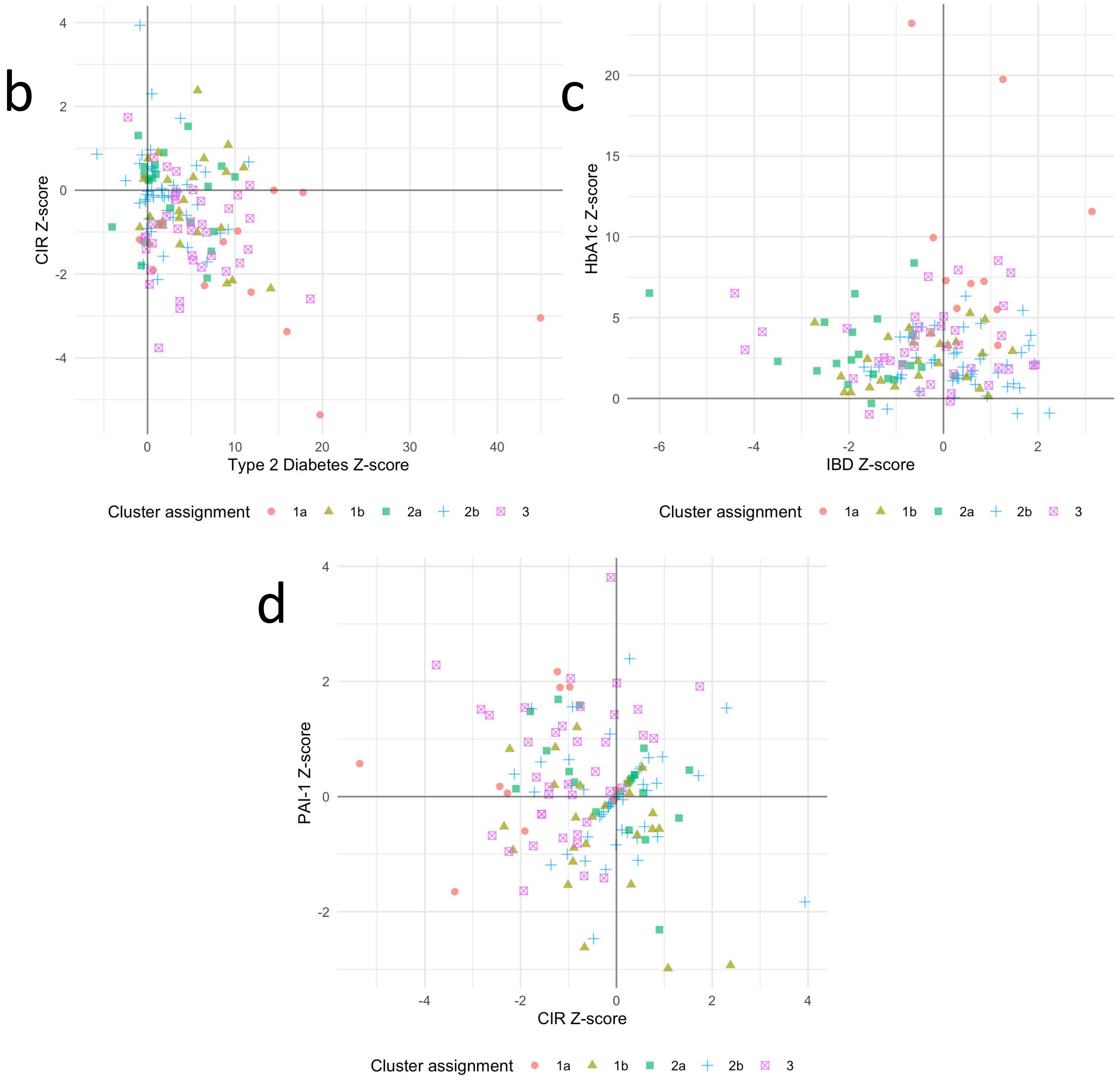
b
CIR Z-score
Type 2 Diabetes Z-score
Cluster assignment 1a 1b 2a 2b 3
c
HbA1c Z-score
IBD Z-score
Cluster assignment 1a 1b 2a 2b 3
d
PAI-1 Z-score
CIR Z-score
Cluster assignment 1a 1b 2a 2b 3
